# Belantamab Mafodotin Triggers Immune Invigoration in Multiple Myeloma Via Inflammatory and Immunogenic Cell Death

**DOI:** 10.1101/2025.09.11.25335116

**Authors:** Edmund CR Watson, Halima Ali Shuwa, Morgan Heycock, Jenny Wade, Chen-Yi Wang, Mint Htun, Lingzi Li, Vivianne Niehaus, Warren Baker, Benjamin Miller, Ruben M Drews, Paul R Barber, Yinjiao Ma, Diana Munera, Qingqing Hong, Daniel E. Lowther, Ali Cenk Aksu, Lydia Lee, H. Christian Eberl, Eleanor Calcutt, Sarah Gooding, Yuqi Shen, Abdullah Khan, Joanna Hester, Fadi Issa, Giorgio Napolitani, Hanny Musa, Karthik Ramasamy, Udo Oppermann, Sue Griffin

**Author notes:** **Correspondence:** Udo Oppermann, Botnar Research Centre, NIHR BRC Oxford, Nuffield Department of Orthopaedics, Rheumatology and Musculoskeletal Sciences, University of Oxford, OX3 7LD, UK; Oxford Translational Myeloma Centre, University of Oxford, OX3 7LD, UK Sue Griffin, GSK, Stevenage, UK SG1 2NY. These authors contributed equally to this work (Udo Oppermann, Sue Griffin).

## Abstract

Belantamab mafodotin, an antibody-drug conjugate targeting B-cell maturation antigen, has demonstrated significant clinical efficacy in combination therapies for relapsed/refractory multiple myeloma. Belantamab mafodotin exerts therapeutic effects through cytotoxicity of its payload, monomethyl auristatin F, and through mediation of antibody-induced cell death. Long-term clinical responses were observed with monotherapy treatment, despite dose holds, suggesting adaptive immune system involvement. Here, we show that belantamab mafodotin induces markers of immunogenic and inflammatory cell death in vitro and ex vivo. Belantamab mafodotin monotherapy treatment triggers acute inflammation detectable in patient serum within 24 hours, with increases in granzyme B, CXCL9, CCL3, and CCL4 linked to response depth achieved. High expression of LRP1 and TLR2, receptors that mediate immunogenic cell death on patients’ monocytoid (monocyte/macrophage) cells, suggests an important function of monocytoid cells to mediate the inflammation and immunogenic cell death cascades. Inflammation is followed by remodeling of the innate and adaptive immune system, a reduction in immune inhibitory signaling and the emergence of CD4 granzyme B-expressing cells in patients in remission vs those that relapse. Belantamab mafodotin’s ability to promote adaptive immune responses and its cytotoxic activity may help explain the durable responses observed in treated patients, despite dose and schedule modifications.

**Graphical abstract:** 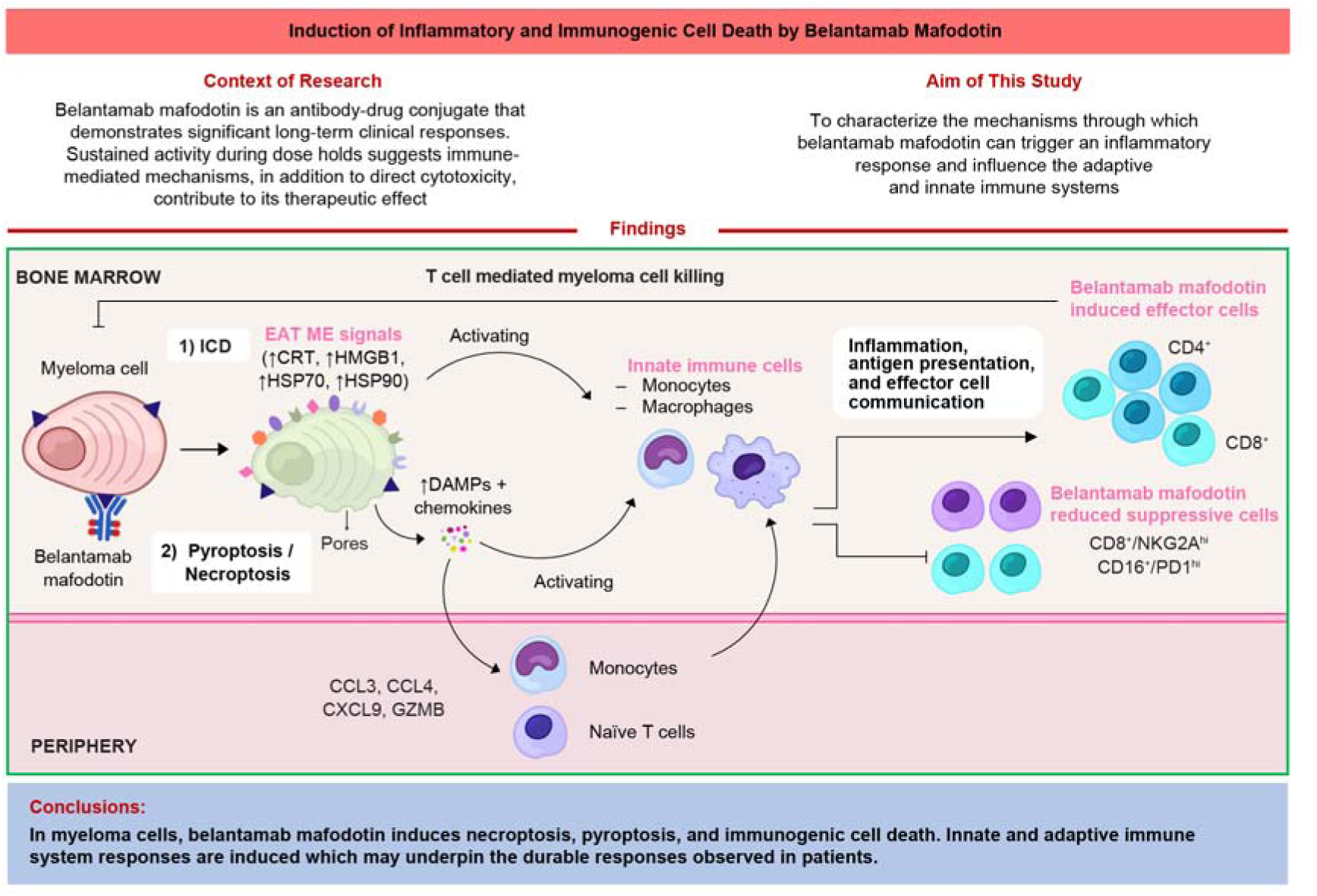

## Introduction

Patients with multiple myeloma (MM), a cancer of bone marrow (BM)-resident plasma cells (PC), typically experience a relapsing and remitting disease course.^1, 2^ The PC BM niche constitutes a tumor microenvironment (TME) densely populated with innate and adaptive immune cells in a complex stromal milieu. MM progression from precursor forms to clinically evident disease occurs alongside TME changes that support MM growth and persistence.^3^

Belantamab mafodotin comprises an afucosylated, humanized anti-B cell maturation antigen (BCMA) monoclonal IgGk1 linked to monomethyl auristatin F (MMAF), an anti-tubulin toxin payload.^4^ The clinical efficacy of belantamab mafodotin monotherapy was initially demonstrated in late-line relapse, where despite modest overall response rates and median progression-free survival (PFS), robust and significant durations of response (DoR) were seen.^5–7^ In phase 3 trials, belantamab mafodotin combinations achieved significant increases in PFS (DREAMM-7 [NCT04246047] and DREAMM-8 [NCT04484623]) and overall survival (OS) (DREAMM-7 [follow-up for DREAMM-8 ongoing]) versus standard-of-care triplets.^8, 9^ Dose/schedule modifications to manage ocular events were frequent in belantamab mafodotin trials.^5, 6, 10^ With monotherapy, responders experienced more dose interruptions than non-responders, ^6^ suggesting that clinical response continues in the absence of active treatment.

Preclinical studies suggest that belantamab mafodotin acts through a combination of natural killer (NK) cell mediated antibody-dependent cellular cytotoxicity (ADCC); opsonization of PCs enabling antibody-dependent phagocytic cytotoxicity (ADCP); and cell death mediated directly by MMAF.^4, 11, 12^ The latter leads to immunogenic cell death (ICD) that could trigger an adaptive immune response and immunologic memory.^4, 11^

ICD is a regulated cell death that features pre-apoptotic surface exposure of calreticulin and the release of ATP throughout cell death and HMGB1 post-mortem.^13^ Release of heat shock proteins and damage-associated molecular patterns (DAMPs) and other factors can accompany ICD.^14^ These “eat me” and “find me” signals trigger dendritic cell (DC) activation via calreticulin and LRP1 (CD91) interaction on DCs,^14^ leading to T-cell-mediated immunologic memory against the tumor cells.^15^ Evidence for belantamab mafodotin-associated ICD is derived from cell line data and a model of murine lymphoma overexpressing BCMA.^11^

While ICD is defined by its ability to promote an adaptive immune response, other forms of regulated cell death are characterized by inflammation alone. Pyroptosis and necroptosis exhibit release of inflammatory chemokines, cytokines, and DAMPs through cell membrane pore formation, leading to local inflammation.^16^ To delineate their activity from classic ICD, we describe these two pathways as inflammatory cell death,^17, 18^ which in some cases, can be triggered by tubulin stabilising toxins.^19, 20^

Here, we demonstrate that belantamab mafodotin, via its anti-tubulin toxin payload, triggers both inflammatory and immunogenic cell death in cell lines, ex vivo in samples from patients with MM and in patients treated with belantamab mafodotin. We suggest that belantamab mafodotin is associated with inflammation and immune priming in successfully treated patients, which contributes to the development of immunologic memory.

## Methods

### Drug treatment of human MM cell lines (HMCLs) and BM mononuclear cells (BMMCs)

HMCLs/ BMMCs were seeded into 96-well plates at 1×10^5^ cells/well and incubated for 48 or 72 hours (HMCLs) or 16 hours (BMMCs) with indicated concentrations of belantamab mafodotin (GSK) or MMAF (Selleck Chem).

### Patient samples

Prior to UK marketing authorization, the named patient program (NPP) permitted belantamab mafodotin monotherapy to be available to patients who had no suitable alternative treatment options. Additional belantamab mafodotin treatment samples were obtained from the DREAMM-14 monotherapy study (NCT05064358), or the DREAMM-5 platform study (NCT04126200)^21^ where indicated.

### Flow cytometry

Briefly, HMCL or BMMCs were washed with phosphate-buffered saline (PBS) and resuspended in live/dead stain (1:400 dilution, Biolegend) and FcR block solution (1:100 dilution; Miltenyi) for 20 minutes before pelleting. Cells were stained with conjugated antibodies for 60 minutes, fixed, permeabilized, stained with intracellular antibodies for 60 minutes, followed by secondary antibodies for 40 minutes and acquired on a Cytek Aurora. In BMMC samples, viable MM BCMA+ cells were defined as CD45-CD40+BCMA+. Data were analyzed using FlowJo v10.10.0. Statistical analysis was performed in R 4.4.1. For each marker, a binomial proportions model was used and the Benjamini-Hochberg (BH) multiplicity correction was applied to control the false discovery rate (FDR) at 5%.

Longitudinal peripheral blood mononuclear cell (PBMC) samples from 33 patients in the ongoing DREAMM-5 study^21^ were analyzed by flow cytometry using a TNBK immune cell phenotyping panel. Statistical analysis was performed using the Linear Mixed Model (LMM) to model the longitudinal data [lmer()] function from the lme4 package in R.

### Bulk RNA sequencing

RNA was extracted using a KingFisher Apex system according to manufacturer’s protocol and strand-specific bulk RNA sequencing conducted (RNAseq) (poly-A selection, Illumina 2×150 bp paired-end sequencing, 100 million reads). Differential gene expression analysis was performed using DESeq2.^22^ Pathways were identified in Reactome DB and tested for self-contained gene sets correcting using fry from the edgeR package. Log2 fold changes between control and test agent were moderated using ashr.^23^ Multiple testing correction was performed by controlling the FDR using Independent Hypothesis Weighting.^24^

### Chemokine bioassay

Cell supernatant or serum from 43 patients from DREAMM-14 treated with belantamab mafodotin monotherapy were evaluated using a U-plex custom 10-plex kit (Mesoscale Discovery Platform; MSD) or HMGB1 ELISA (Cloud Clone) according to manufacturer’s instructions. Concentrations were calculated using a standard curve for each analyte and corrected for dilution factor. Statistical analysis was performed in R version 4.4.1. For each secreted inflammatory mediator, a Linear Mixed or Bayesian Tobit model was implemented. Best confirmed response was used to determine response categories (Supplemental Table 6).

### Proteomics

NCI-H929 (ATCC) supernatant was analyzed using Olink Explore 1536 according to manufacturer’s instructions. Libraries were sequenced on a NovaSeq 6000 (Illumina). The lower limit of detection (LOD) provided by the NPX Explore software (v.3.9.0) was utilized for censoring. Statistical analyses were conducted using R version 4.3.1, with linear models fitted separately to each protein using the lm() function. P-values were adjusted for multiple testing using the Benjamini-Hochberg FDR method.

### Multiplex immunofluorescence imaging

Twenty formalin-fixed paraffin-embedded trephine BM biopsies from 10 patients from DREAMM-5 sectioned to 4 µm were mounted on SuperFrost Ultra Plus™ GOLD Adhesion Slides (Epredia). Slides were baked, dewaxed, and treated with BOND Epitope Retrieval Solution 2 (Leica) using Leica BOND RX auto stainer. Imaging was performed using the Leica Cell DIVE system through 7 rounds of fluorescent-label antibody staining, imaging, and dye inactivation with 3% hydrogen peroxide (Merck) and 0.1 M sodium bicarbonate (Merck). Image analysis used custom Python scripting. MM/PC were classified as CD138+BCMA+. Generalized least squares models were used to assess the percent cell type, density of cell type, or biomarker intensity change.

### BM samples from belantamab mafodotin-monotherapy treated patients

BM aspirates acquired as part of routine care underwent CD138 negative selection (StemCell EasySep) and the CD138- fraction was separated on a Ficoll gradient and viably frozen.

### Mass cytometry

Cells (3×10^6^ per sample) were washed twice in Maxpar Cell Staining Buffer (CSB; Standard BioTools) and stained with pre-aliquoted surface and intracellular antibody cocktails (Supplemental Table 5). Data were acquired on a Helios mass cytometer (Standard Biotools), according to the manufacturer’s protocol.

### Single-cell transcriptomics

BM aspirate cells were processed using GEM-X beads (10X Genomics), with cDNA library amplification per manufacturer’s protocol (10X Genomics, user guide CG000731). cDNA libraries were quantified with TapeStation and 10 ng cDNA taken forward into the Oxford Nanopore (ONT) protocol using PCS114 chemistry on a Promethion P24 using R10.4.1 flow cells (ONT). Super-high accuracy basecalling was performed by Dorado within the MinKNOW software. FASTQ files were processed by running a singularity image of the wf-single-cell pipeline (EPI2ME) on a high-performance cluster, with the reference genome GRCh38 and expected cells set to 5000. Resulting transcript and counts matrices were imported into a Python environment and analyzed using a combination of scanpy,^25^ SoupX,^26^ scDblFinder,^27^ and scVI.^28^ Barcodes with counts and transcripts >5 median absolute deviations below the median and containing ≥20% mitochondrial genes were filtered.

### Spatial transcriptomics

BM trephine sections (5 µm-thick) were assessed by 480-gene custom Xenium panel (Supplemental File 1). Slides were processed on 10X Xenium v1 (10X Genomics) including cell segmentation staining (10X Genomics, user guide CG000749, Rev A). Counts matrices and associated centroid and segmentation coordinates were read into Seurat for R.^29^ Following clustering and manual cell annotation, data were tiled with 500 µm × 500 µm squares and clustered according to cell composition. CellChat analysis was performed to assess communication per tile.^30^

All statistical analyses for biomarker data was performed in a post-hoc, exploratory manner.

## Results

### Belantamab mafodotin response can persist through prolonged dose holds

In patients treated with belantamab mafodotin monotherapy as part of the NPP, 3-month gaps between doses (where the labelled dosing schedule was once every 3 weeks [Q3W]) could be associated with ongoing remission (Supplemental Figure 1), even with clinical responses of less than complete remission (CR). We hypothesized that ongoing responses in the absence of active treatment might be enabled by changes in the TME triggered by belantamab mafodotin.

### Belantamab mafodotin triggers inflammatory and immunogenic death in MM cell lines and patient BM ex vivo

Mechanisms of belantamab mafodotin-induced cell death were characterized to explore how belantamab mafodotin affects the TME. Treatment of HMCLs with belantamab mafodotin triggered significant, dose-dependent, upregulation of ICD markers (calreticulin, HMGB1, HSP70, and HSP90) on the cell surface as well as upregulation of pore-forming proteins involved in the inflammatory cell death pathways, necroptosis (MLKL) and pyroptosis (GSDME and GSDMD) (Figure 1A and Supplemental Figure 2A). These findings were recapitulated with free MMAF, indicating these pathways are induced by the payload (Figure 1B). RNAseq of NCI-H929 cells treated with 10 μg/mL belantamab mafodotin identified upregulation of necroptosis (p=2.5×10^-11^) and pyroptosis (p=1.8×10^-4^) pathways based on a self-contained test of Reactome gene sets (Figure 1C–D and Supplemental Figure 2B).

**Figure 1.**
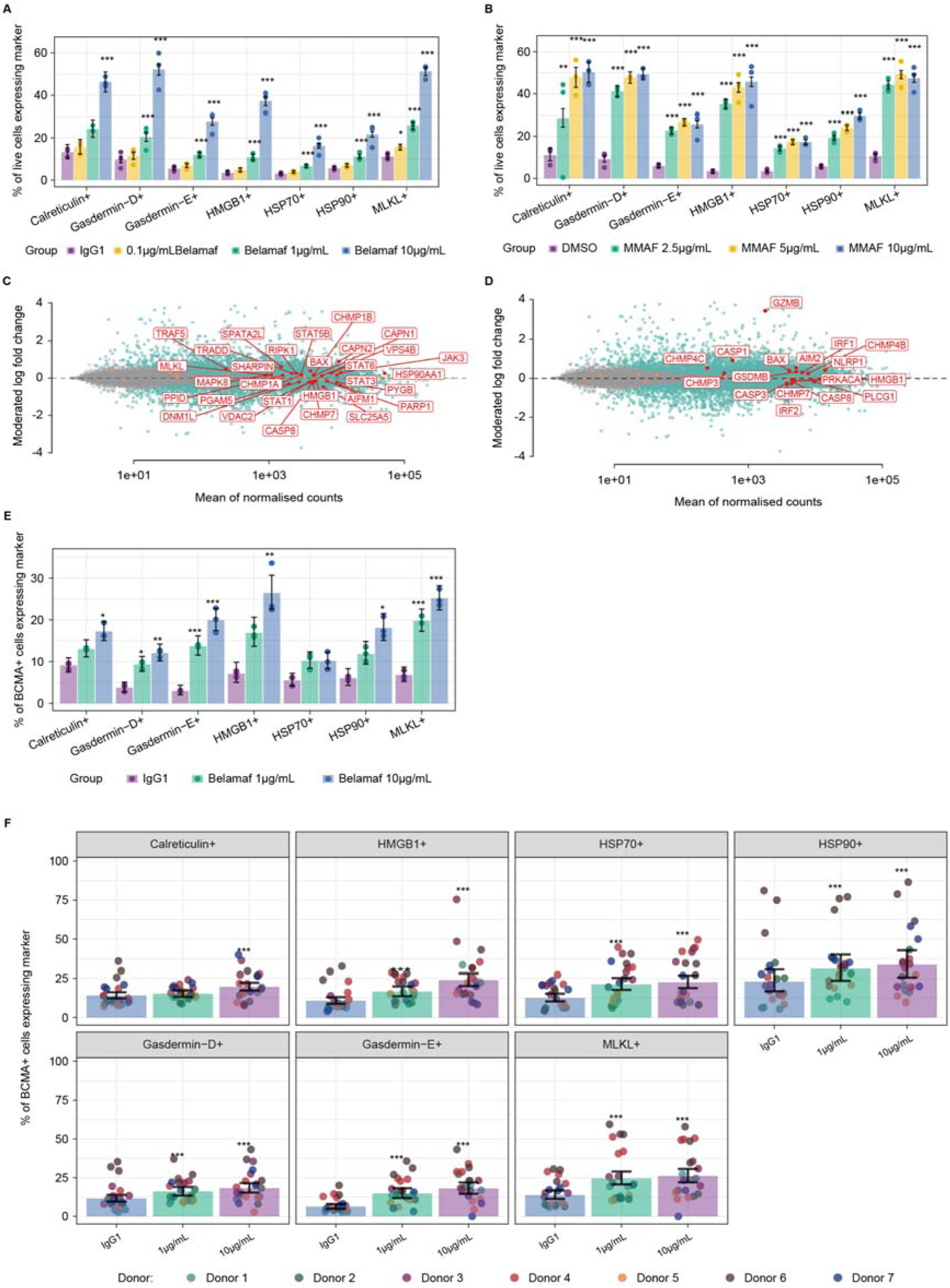
Belantamab mafodotin triggers hallmarks of ICD and inflammatory cell death in NCI-H929 and primary MM cells Percentage of NCI-H929 cells expressing distinct markers of ICD at the cell surface as measured by spectral flow cytometry after 72 hours of treatment with various concentrations of belantamab mafodotin (belamaf) or IgG1 as a negative control (A) or free-MMAF with DMSO as a negative control (B). Data representative of n=3 independent repeats. (C–D) MA plot of RNAseq results of belantamab mafodotin 10 µg/mL vs IgG control showing mean log2 expression on x-axis and moderated log2 fold change on y-axis. FDR was controlled using Independent Hypothesis Weighting, with genes meeting the significance threshold (FDR < 0.1) are shown in green. Genes implicated in cell death pathways are highlighted in red for necroptosis (C) and pyroptosis (D). RNAseq data representative of that obtained from 2x different cell lines. (E–F) Primary patient BMMCs were incubated with belantamab mafodotin for 16 hours and cell surface markers were assessed by spectral flow cytometry. For each marker, a binomial proportions model was used to compare treatment response vs IgG1 vehicle control and the BH multiplicity correction was applied to control the FDR set at 5%. Graphs represent an exemplar plot from an individual donor (E) and amalgamated data from n=7 donors where the colours represent donors and the dots represent replicates (F). Error bars are standard error of the mean, ***P<0.001; **P<0.01; *P<0.05. Belamaf, belantamab mafodotin.

We next treated BM mononuclear cells (BMMCs) from 7 MM patients ex vivo for 16 hours. Flow cytometry analysis confirmed that belantamab mafodotin and MMAF induced significant upregulation of all tested markers in BCMA-positive PCs, validating the cell line findings (Figure 1E-F and Supplemental Figure 2C). BMMCs were treated with daratumumab as a negative control, which did not induce markers of ICD, necroptosis, or classical pyroptosis (GSDMD); GSDME increased, as expected, since BMMCs contain NK cells, and GSDME expression can be triggered by NK cell killing via ADCC,^31^ a known mechanism of action of daratumumab (Supplemental Figure 2D).

Pyroptosis- and necroptosis-associated pores enable the release of inflammatory chemokines and cytokines. We used Olink-based proteomics to probe for proteins in the supernatant of NCI-H929 cells incubated with belantamab mafodotin and MMAF. Of 1432 measured proteins, >100 were significantly elevated following 10 μg/mL belantamab mafodotin treatment, consistent with inflammatory cell death (Figure 2A, Supplemental Figure 3A and Supplemental File 2). Focusing on immune modulators, belantamab mafodotin and MMAF treatment resulted in accumulation of granzyme B (GZMB) and the proinflammatory mediators CXCL8 (IL8), CCL3, and CCL4, which are crucial for immune cell recruitment to inflamed tissues. GZMB is typically associated with cytotoxic T/NK cells; therefore, elevated secretion of this marker from MM cells in response to belantamab mafodotin was unexpected. However, all these factors are NF-kB response genes, which is a key downstream module of the necroptotic cell death pathway; additionally, GZMB cleaves GSDME, indicating a potential interconnection between the necroptosis and pyroptosis pathways in belantamab mafodotin-treated cells (Figure 2A).

**Figure 2.**
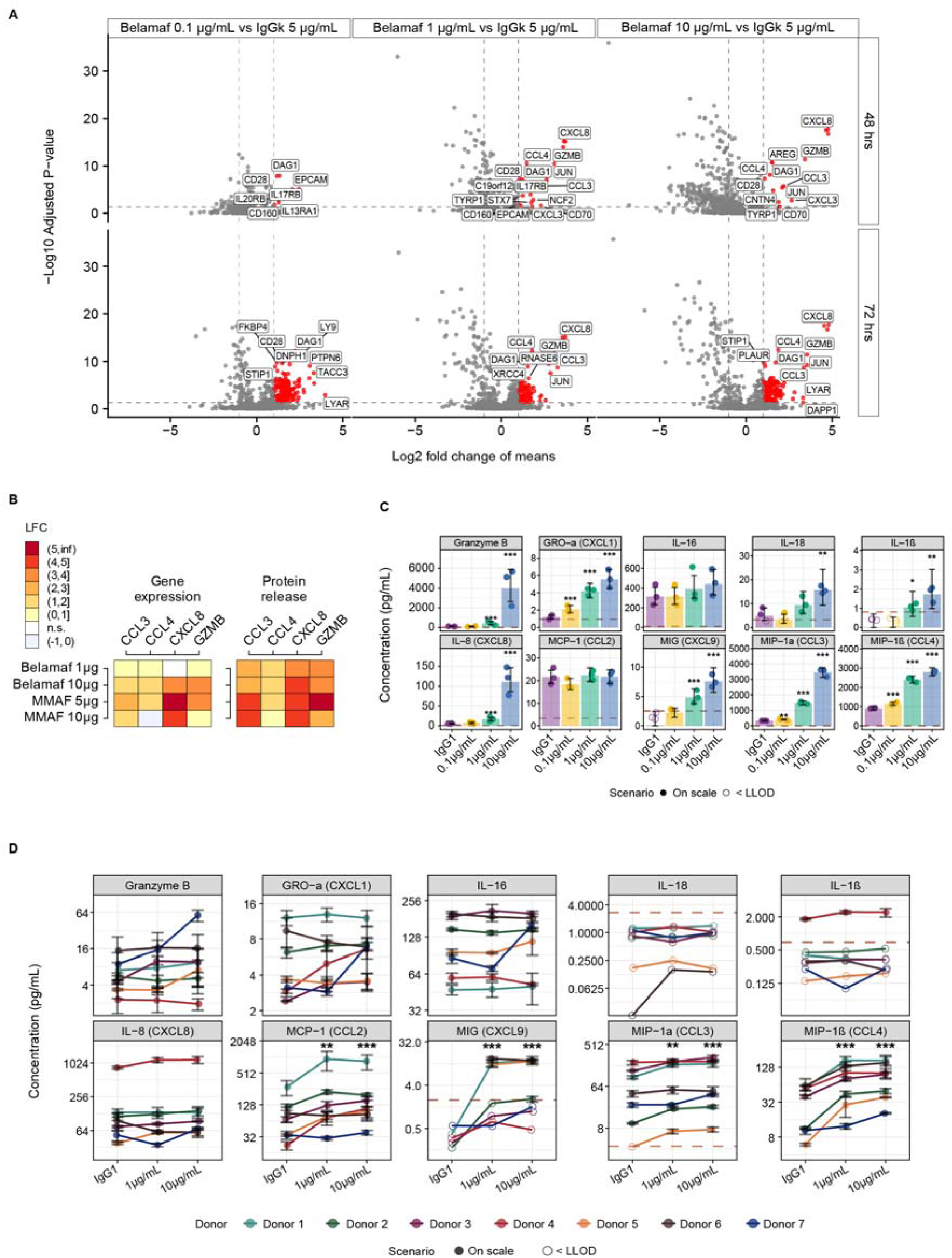
Soluble markers of inflammatory and immunogenic cell death are enriched in the supernatant of belantamab mafodotin-treated NCI-H929 and primary MM cells (A) Volcano plots visualizing the Log2 fold changes in analyte concentrations in the supernatant from NCI-H929 cells treated for 48 or 72 hours with belantamab mafodotin (belamaf) at 0.1, 1, and 10 µg/mL. Proteins showing a Log2 fold change of at least 1 and an adjusted P-value of <0.05 are considered significantly modulated and are colored in red. Selected significant proteins are indicated by gene name. Proteins with decreased expression are not included in the analysis, since it is not possible to delineate true reductions vs reductions due to the death of the treated cells. (B) Heatmap comparing transcript changes (RNAseq, moderated LFC) with secreted protein levels in supernatants from belantamab mafodotin-treated cells (Olink, LFC) for CCL3, CCL4, CXCL8, and GZMB. n.s. = not significant (FDR > 0.05). (C) Concentration of selected pro-inflammatory molecules in the supernatant of NCI-H929 cells treated with belantamab mafodotin as measured by MSD to validate the proteomics findings. Dotted line indicates lower limit of detection (LOD) of the assay in the linear range. Error bars are standard error of the mean. Chemokine validation data is representative of that derived from n=3 independent experiments. (D) Concentration of the same molecules in the supernatant of belantamab mafodotin-treated primary patient cells from n=7 donors. Dotted lines indicate lower limit of detection of the assay in the linear range. Error bars are standard error of the mean. For each of the secreted inflammatory mediators, a Bayesian Tobit model was implemented to compare treatment vs IgG control. ***P<0.001; **P<0.01; *P<0.05. Belamaf, belantamab mafodotin.

RNAseq of NCI-H929 cells showed significantly elevated levels of GZMB, CXCL8, CCL3, and CCL4 following belantamab mafodotin and MMAF treatment, suggesting that the increase of chemokines results from transcriptional reprogramming, rather than simple cytosolic protein ‘leakage’ following cell necrosis (Figure 2B and Supplemental Figure 3B). Using MSD, a high-sensitivity sandwich immunoassay, we additionally tested for previously reported pyroptosis and necroptosis-associated chemokines which were below the limit of detection in the proteomic analysis, and observed increases in CXCL1, IL16, IL18, and IL1β (Figure 2C and Supplemental Figure 3C).

Investigating BMMCs from patients with MM treated ex vivo with belantamab mafodotin for 16 hours confirmed the release of CCL3, CCL4, CCL2, and CXCL9; the secretion of other proinflammatory mediators was not confirmed, likely due to shorter incubation time of primary cells (Figure 2D). The secretion of proinflammatory mediators was not detected with free-MMAF treatment, possibly due to excessive toxicity with primary cells (also noted in NCI-H929 cells, where high MMAF concentrations led to lower secretion) (Supplemental Figure 3C).

Together, these data support simultaneous activation of programmed cell death pathways that are immunogenic and inflammatory, triggering chemokine/cytokine release that could support influx of immune cells into the BM.

### Belantamab mafodotin monotherapy treatment induces a pro-inflammatory chemokine signature in serum of MM patients

To study belantamab mafodotin-triggered inflammation in patients, we analyzed serum from 43 patients, collected at baseline, 24 hours post treatment, and 3 weeks post-dosing (prior to the second treatment cycle) with belantamab mafodotin monotherapy in the DREAMM-14 study.^32^ Immunoassay analysis demonstrated significant spikes in the same pyroptosis- and necroptosis-related inflammatory factors discovered in vitro, 24 hours post first dose (Figure 3A and 3B). These included the ICD-associated DAMP, HMGB1, and inflammatory factors CCL2, CCL3, IL8, IL16, IL1β, CXCL9, GZMB, and CCL4, which returned toward baseline levels prior to the second cycle, highlighting the acute and transient nature of the belantamab mafodotin-triggered inflammatory response.

**Figure 3.**
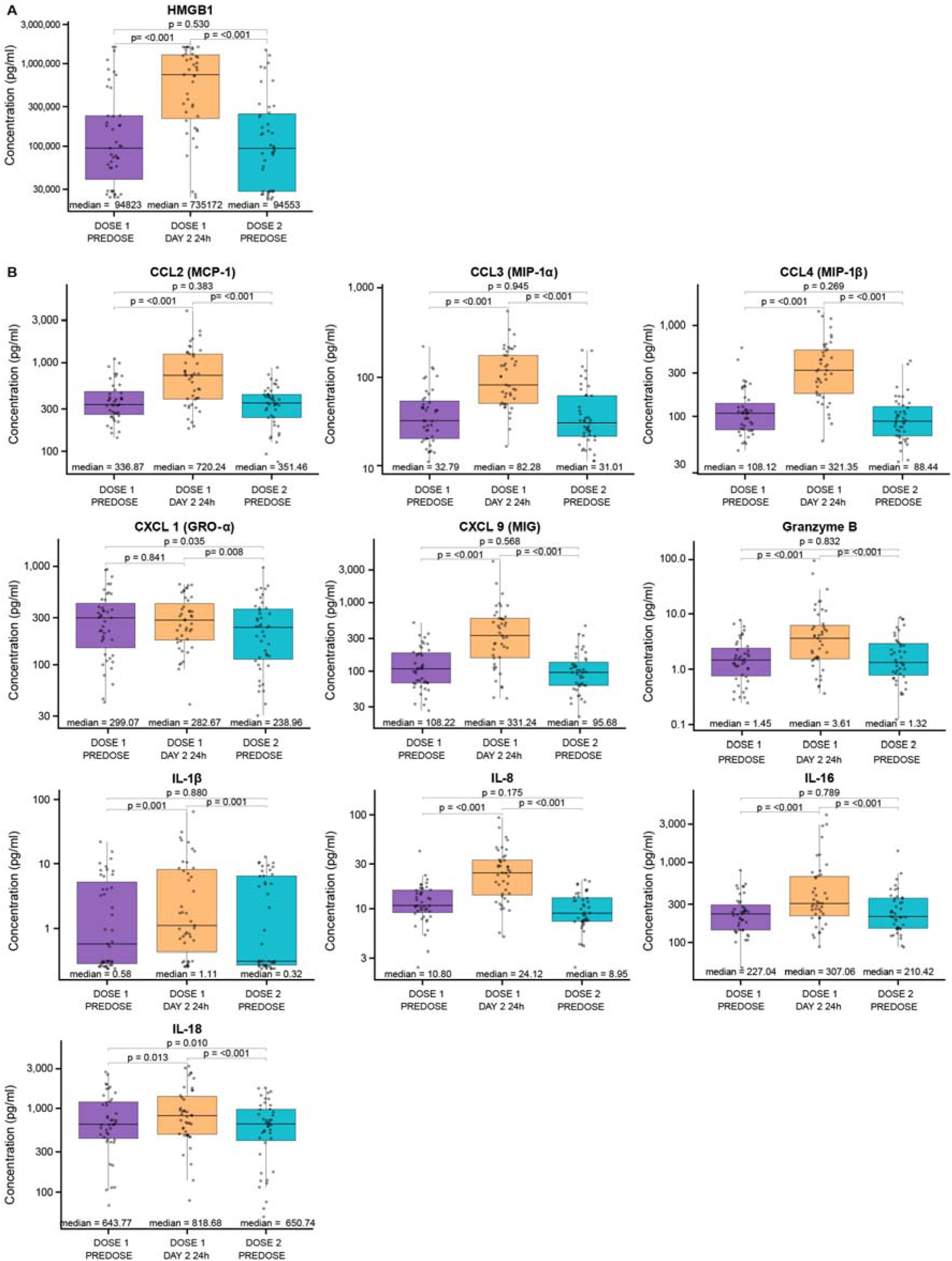
Belantamab mafodotin triggers hallmarks of acute, transient inflammation in patient serum Concentration of inflammatory mediators in serum samples from n=43 patients in DREAMM-14 treated with various dosing regimens of belantamab mafodotin monotherapy. Serum samples were collected at three time points for each patient: Dose 1 – Predose; 24 hours post-dose 1; and prior to Dose 2. Statistical analysis was conducted using a Bayesian Tobit models (for HMGB1 and IL1β markers) or a Linear Mixed-effect Models (LMM) for the remaining markers. The p-values for pairwise comparisons of the three time points obtained from the LMM have been adjusted using the Tukey-Kramer method (A) Quantification of the ICD marker HMGB1 by ELISA. (B) Comparative concentrations of secreted IL8, Granzyme B, IL16, CCL3, CCL4, IL18, IL1β, CXCL9, CCL2, and CXCL1 quantified using the MSD 10-plex assay across different dosing regimens.

We used a linear mixed statistical model to evaluate whether the longitudinal profile of these markers differed according to best clinical response achieved by a patient (time-by-response interaction). GZMB was significantly increased compared to baseline at 24 hours in deep responders (≥ very good partial response) but not in non-responders (progressive disease) (Figure 4).

**Figure 4.**
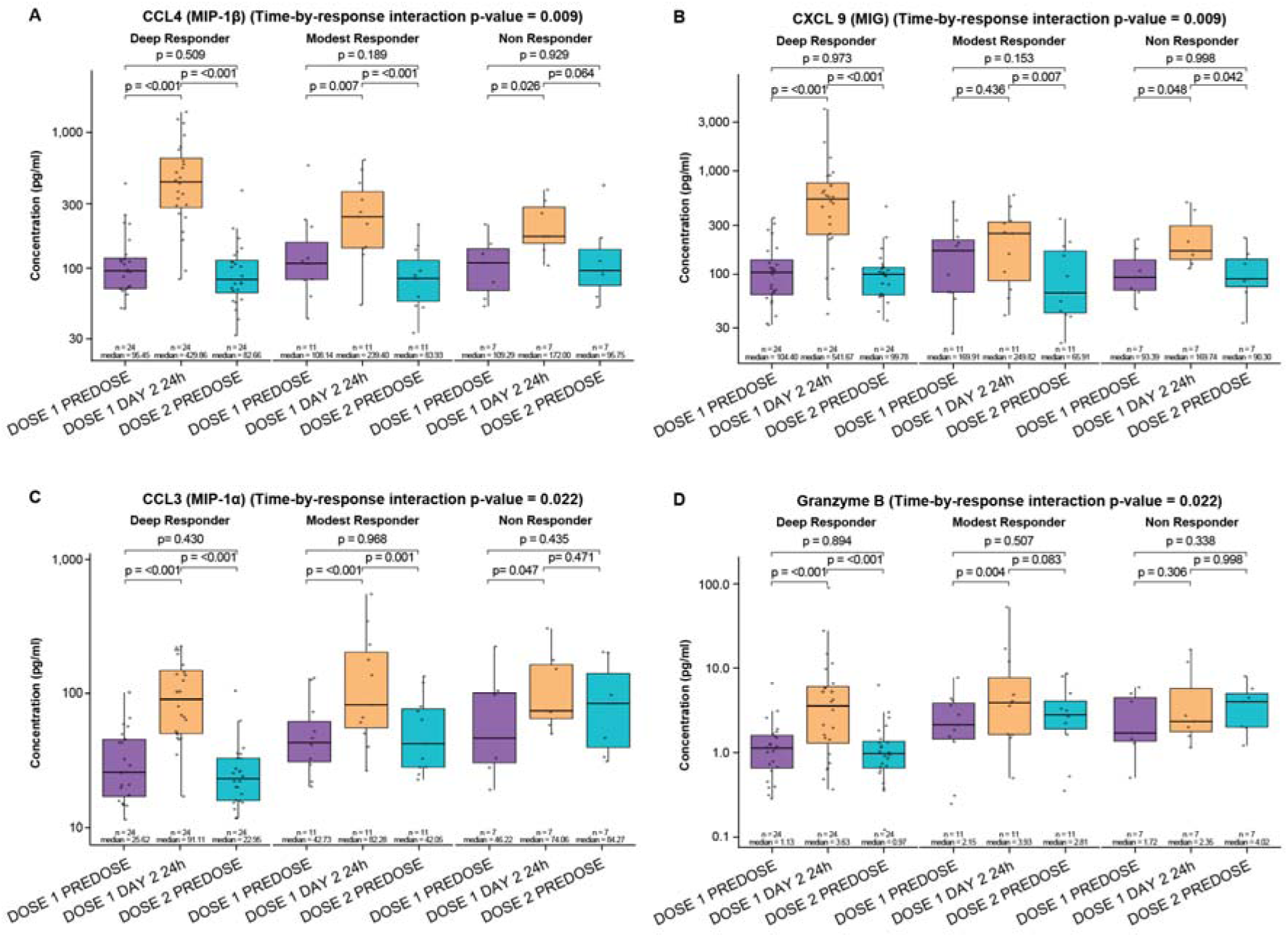
A subset of inflammatory chemokines was associated with the depth of response in patients Concentration of inflammatory mediators (A) CCL4, (B) CXCL9, (C) CCL3 and (D) Granzyme B in serum samples from patients in DREAMM-14 treated with belantamab mafodotin monotherapy. Serum samples were collected at three time points for each patient: Dose 1 – Predose; Dose 1 – 24 hours post-dose; and Dose 2 – Predose. Data are grouped by patient response. Deep responder = patients who achieved a very good partial response (VGPR), complete response (CR), or stringent complete response (sCR). Modest responders = patients who achieved a partial response (PR), minimal response (MR), or stable disease (SD). Non-responders = progressive disease (PD). LMM was used to evaluate if the longitudinal profile of a marker differed by response categories. The overall difference is characterized by the “interaction P-value” which has been adjusted for multiple testing of the 9 markers using the Benjamini-Hochberg FDR method. For pairwise comparisons between three time points within response groups, the p-values have been adjusted using the Tukey-Kramer method.

Additionally, CXCL9, CCL3, and CCL4 were identified through the time-by-response interaction assessment; these markers significantly increased at 24 hours in deep responders, with less pronounced increases in modest responders (i.e., those who achieved a partial response, minimal response, or stable disease) and non-responders. Baseline GZMB and CCL3 levels appeared lower in deep responders vs modest and non-responders, but this difference was not significant. Overall, successful induction of inflammatory chemokines within 24 hours following treatment may trigger downstream immune infiltration and remodelling to support deep responses.

### The BM monocytoid population expresses receptors necessary for mediating ICD

To determine which TME components might receive and elaborate the inflammatory signal from dying PCs, single-cell RNAseq analysis was used to compare BM aspirates from 7 patients before belantamab mafodotin monotherapy treatment (administered as part of the NPP) with control BM samples from 4 age-matched patients (taken from femoral heads in patients undergoing elective hip arthroplasties for non-malignant disease) (Supplemental Figure 1 and Figure 5A-B).

**Figure 5.**
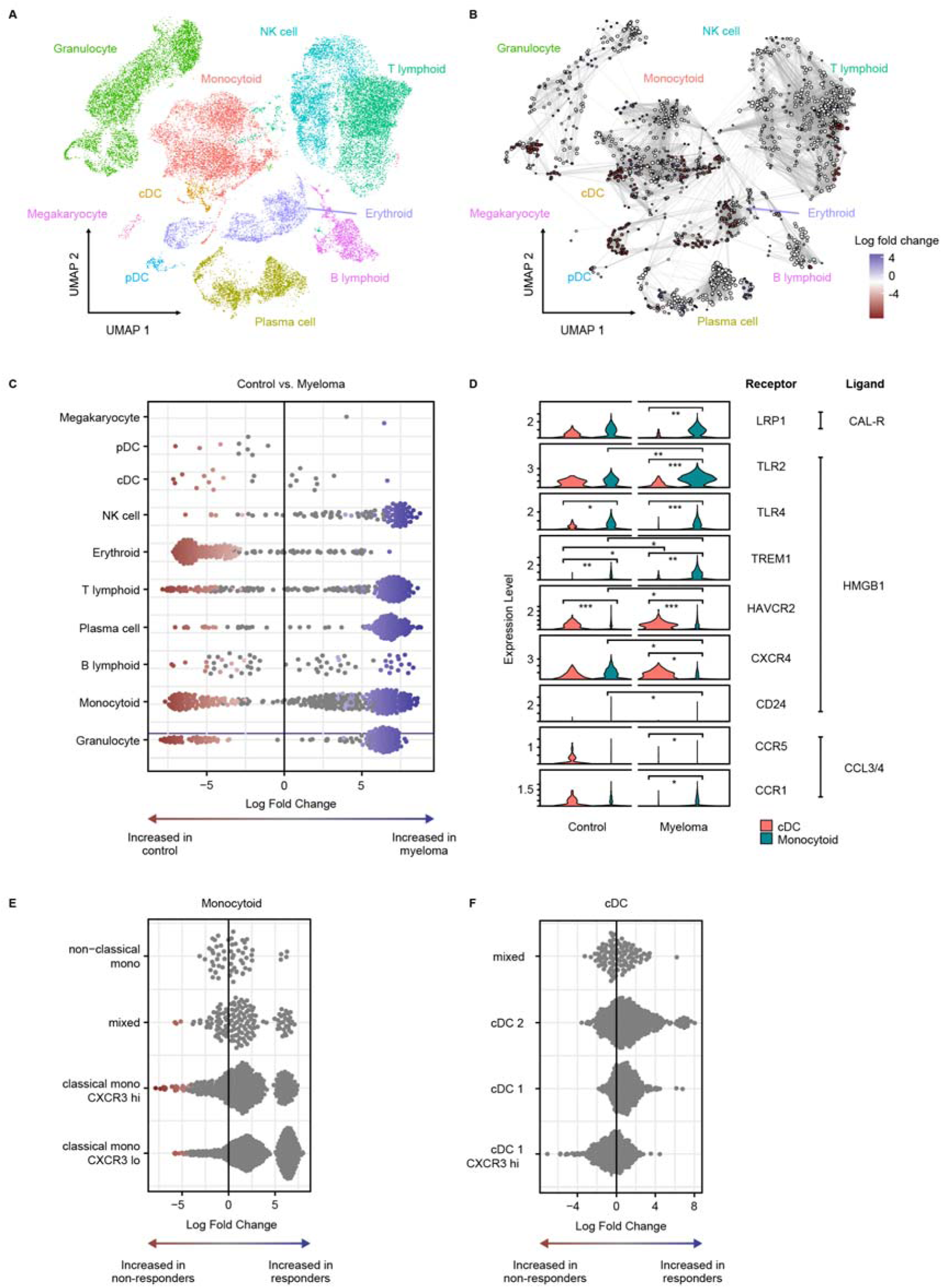
Monocytoid cells are a dominant cell type in the MM TME and are equipped to respond to ICD Samples from patients in the named patient program who received belantamab mafodotin monotherapy: (A) UMAP dimensionality reduction of long read scRNAseq data from patient samples (15 samples total, across 12 separate patients) plus 4 control samples, integrated by scVI and colored by cell type. (B) Neighbourhood analysis of pre-treatment patient samples compared with control samples, plotted in the same UMAP axes as (A), where blue shading in the circles means increased in patient samples and red means increased in control samples. Size of circles proportionate to size of neighborhoods, and thickness of edges related to connectivity of neighborhoods. (C) Beeswarm plot showing differential abundance of cell types as determined by scRNAseq in pre-treatment patient samples vs control; as with (C), more negative numbers suggest more abundant in controls, more positive numbers indicate more abundant in patients. Each point represents a neighbourhood, which are coloured if they are significantly differentially abundant at an FDR of 0.1. (D) Violin plots of scRNAseq data showing relative expression of key immunogenic and inflammatory receptors in monocytoid and cDC populations, grouped according to ligand; long read scRNAseq data. Pseudobulk analysis using DESeq2 method for differential expression. Multiple hypothesis testing adjusted for by Benjamini-Hochberg method, where P-adjusted values for comparisons are * P<0.05, ** P<0.005, *** P<0.0005. Only the receptors with significant results are shown. (E–F) Beeswarm plots of mass cytometry data of the same MM baseline samples, showing differences in the monocytoid (E) and classical dendritic cell (cDC) (F) populations in responding vs non-responding patients at baseline. Positive values represent over-representation in responders. Non-paired analysis, performed with MiloR, where colored dots refer to statistically significant events at the FDR value of 0.2.

Baseline (pre-belantamab mafodotin initiation) samples demonstrated expanded T-lymphoid and NK cell compartments in patients with MM relative to non-malignant controls according to differential abundance analysis (Figure 5B-C). Amongst innate immune cells, the MM TME had fewer classical and plasmacytoid dendritic cells, more granulocytes, and in some cases, more monocytoid (monocyte/macrophage) cells. Classical DCs (cDCs) are canonical mediators of ICD, yet expression of pro-inflammatory ICD receptors LRP1, TLR2, TLR4, and TREM1 was significantly greater in MM monocytoid cells compared to cDCs. This appears to be an MM-specific observation, because no such (or less appreciable) differences were observed in control samples (Figure 5D). Of interest, the inhibitory ICD receptor, TIM-3,^33^ encoded by HAVCR2, is much more highly expressed in cDCs than monocytoid cells, in both control and MM patients. TLR2, TREM1, HAVCR2, and CXCR4 were differentially expressed in MM vs control monocytoid cells, suggesting MM-specific modulation of the monocytoid compartment (Figure 5D).

We applied a 39-parameter mass cytometry panel to the same samples. Differential abundance analysis of the baseline state by MiloR showed a non-significant increased representation of monocytoid populations, but not cDC, in responders compared to non-responders (Figure 5E-F). Additionally, the observation that monocytoid chemokines (CCL3 and CCL4) transiently increase in the serum of deep responders (Figure 4) indicates a potentially important contribution of this cell type to therapy response. Together, these data indicate monocytoid cells are the most likely myeloid population involved in relaying the immunogenic and inflammatory signal emitted by belantamab mafodotin-induced PC cell death.

### Successful belantamab mafodotin treatment is associated with reduction in immunosuppressive communication and emergence of a key immune effector population

We sought changes in the TME that could represent sequelae of a belantamab mafodotin-triggered, monocytoid-amplified, acute inflammatory state. In situ spatial transcriptomics assessed cell–cell communication networks in matched baseline/remission samples from a responding patient, and additionally in a follow-up sample from a patient relapsing on treatment (Figure 6A-B). PC proximity was controlled by tiling over each trephine and selecting tiles of approximately equal cell composition for comparison (Supplemental Figure 4A). We found a reduction in the inhibitory signaling axes (MIF and LGALS9) from baseline to remission, with higher levels in relapse (Figure 6B and Supplemental Figure 4B-D); the T- and NK-cell exhaustion marker, TIGIT showed a similar pattern (Figure 6B). These communication changes from 2 patients were consistent with a broader trend of immune de-repression and invigoration seen across our mass cytometry dataset. Here, 5 responding patients had a larger, putatively inhibitory, CD16 and PDL1-double positive granulocyte population at baseline that diminished over time compared with the 2 non-responders (Figure 6C and 6D).

**Figure 6.**
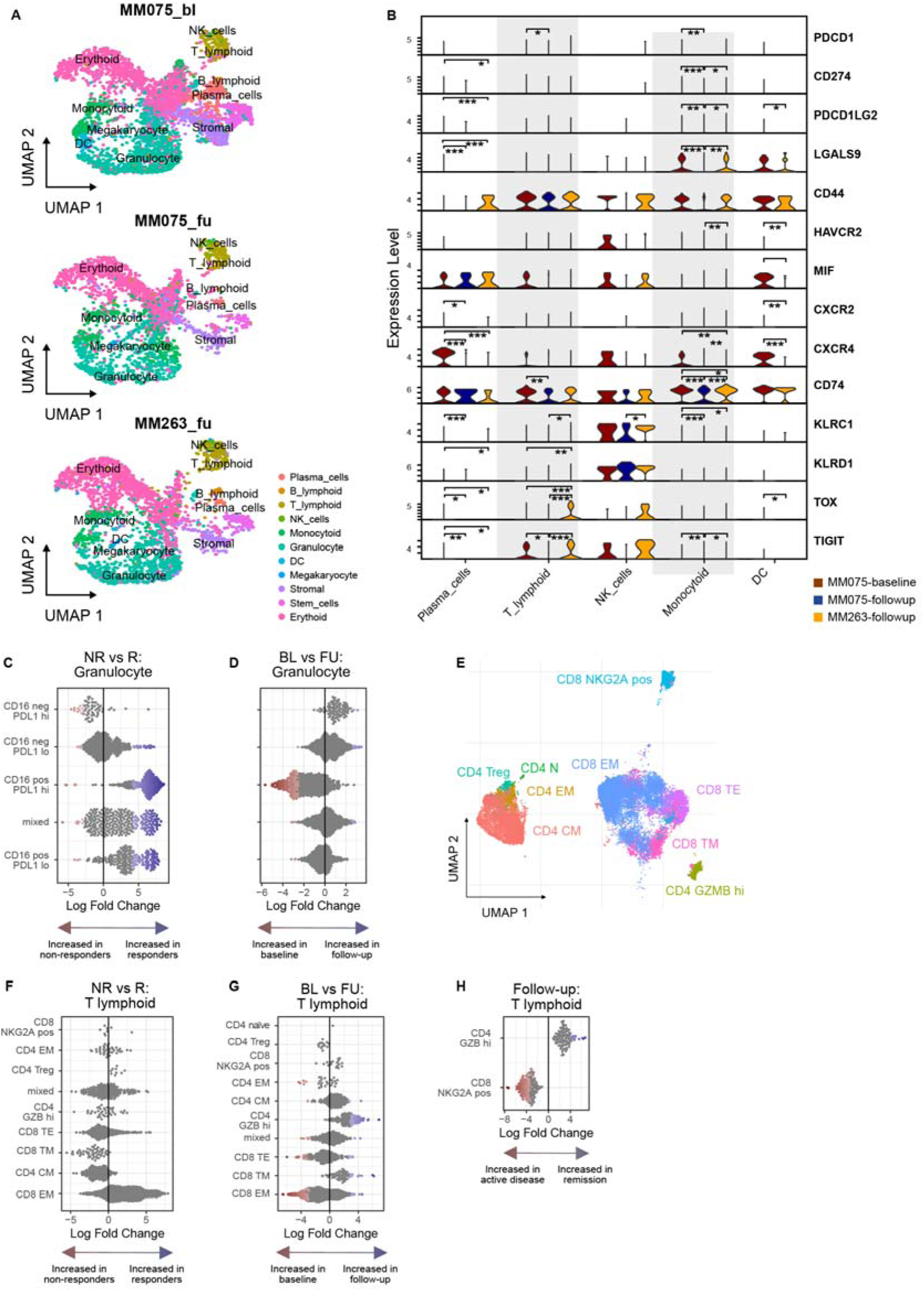
The BM of responding patients shows signs of invigoration and emergence of adaptive immunity Samples from patients in the named patient program who received belantamab mafodotin monotherapy: single cell in situ spatial transcriptomics was performed on three MM BM trephines (see Supplemental Figure 4). (A) UMAP plots of integrated spatial transcriptomic data from selected tiles across 3 patient samples: patient MM075 baseline (top); patient MM075 remission (middle); patient MM263 relapse (bottom). (B) Violin plot of selected immune checkpoint and exhaustion markers, reflecting expression levels in tiles selected for analysis. Statistical comparisons were performed using Seurat FindMarkers with t-test as test model. Significance is indicated as follows: BH-adjusted P<0.05 (*), P<0.005 (**), P<0.0005 (***). (C–H) Mass cytometry data from full monotherapy cohort. (C–D) Beeswarm plots of granulocytes showing changes in cellular neighbourhoods in responders vs non-responders at baseline (C) (where positive values represent over-representation in responders) and in baseline vs follow-up in responders (D) (where positive values represent over-expression at follow-up). (E) UMAP of mass cytometry data of T-lymphoid cells only. (F–G) Beeswarm plots of T cells analyzed as per granulocytes, comparing responders vs non-responders at baseline (F) and baseline vs follow-up in responders (G). (H) CD4 GZMB high cells and CD8 NKG2A high cells analyzed at follow-up, where more positive values indicate greater abundance in the remission state. All beeswarm plots produced by non-paired analysis in MiloR, with neighborhoods shaded if significant at an FDR of 0.2.

Belantamab mafodotin-associated inflammation and immune de-repression may link to an emergent functional adaptive immune response. In the monotherapy cohort, the CD8 effector memory (CD8 EM) compartment in the baseline BM was non-significantly expanded in responders versus non-responders, and reduced in responding patients on treatment, while the CD4 central memory (CD4 CM) compartment appeared to expand (Figure 6F and 6G). These observations were corroborated in an independent cohort of patients from DREAMM-5 (a platform study assessing various belantamab mafodotin combination regimens; details in Supplemental Materials), where we performed multiplex immunofluorescent imaging of BM trephines (Supplemental Figure 5A–B). There was an increase in the ICD marker HMGB1 and a trend towards expansion of CD4 and CD8 CM in responders with a decrease in CD4 and CD8 EM (Supplemental Figure 5C–D). Aligned with the reduced inhibition concept, there was also reduction of an inhibitory (CD11b+) macrophage population in responders (Supplemental Figure 5E). The observed loss of EM in the BM in both cohorts is suggestive of mobilization of functional EMs into the periphery. This concept was confirmed when we assessed the peripheral blood (PB) from patients in the DREAMM-5 study where we observed a significant increase in both CD8 effector and CD8 EM in the periphery following treatment, alongside a significant reduction in terminally exhausted/dysfunctional (TEMRA) T cells—changes consistent with the induction of a functional adaptive immunity (Supplemental Figure 6).

Deeper analysis of the adaptive immune compartment using the mass cytometry data described in Figure 5E-F, showed emergence of a segregated CD4 GZMB-positive population over time in patients treated with belantamab mafodotin monotherapy (Figures 6E-G). At follow-up, this population was significantly more abundant in the remission than the relapse state (Figure 6H). In this group of cells, GZMB levels were equivalent to the CD8 effector populations by normalized mass cytometry expression values (Supplemental Figure 7), consistent with it being a bona fide cytotoxic, MHC-II-restricted effector population.

Collectively, we found changes in TME cellular architecture and communication that are consistent with activation of the adaptive immune system, and correspond with successful belantamab mafodotin therapy.

## Discussion

Enhanced understanding of belantamab mafodotin’s mechanism of action may provide a mechanistic explanation of the robust efficacy observed across clinical trials, including the long-term DoR. Clinically, 3-month gaps between belantamab mafodotin monotherapy doses (where the labeled dosing schedule was Q3W) could be associated with ongoing remission, even when response was less than CR. We hypothesize this might be explained by a model where belantamab mafodotin-induced inflammatory death of PCs stimulates, and is amplified by, monocytoid cells, which then act as an intermediary in the de-repression and invigoration of the TME to induce durable therapeutic response. Whilst our data focuses on belantamab mafodotin, it is important to highlight that these immune-modulatory effects could be complementary to those induced by other agents (e.g., bortezomib and pomalidomide^34, 35^). Accordingly, a recent post hoc analysis of the phase 3 DREAMM-7 and DREAMM-8 trials showed sustained efficacy of belantamab mafodotin in combination with these agents, despite extended dose delays.^36^

Our results suggest that, in addition to ICD, belantamab mafodotin triggers inflammatory cell death by necroptosis and pyroptosis—forms of regulated cell death that release chemokines and DAMPs to trigger innate and adaptive immunity. We observed that the ICD marker HMGB1, alongside pyroptosis and necroptosis-associated inflammatory chemokines, increased in the peripheral serum of patients 24 hours after belantamab mafodotin treatment. While the observed release of HMGB1 is consistent with prior findings,^37^ the increase in other inflammatory mediators associated with cell death represent a novel insight. Increases were seen in: CCL2, CCL3, CCL4, and IL8, which recruit and activate innate cells such as neutrophils and monocytes^38^; CXCL9 and IL16, which recruit T lymphocytes; and IL1β and IL18, which can enhance antigen-specific T-cell responses.^39^ Accordingly, the cell death phenotype triggered by belantamab mafodotin provides an inflammatory milieu that could recruit innate and adaptive immune mediators.^40^ Notably, several of these factors were linked with the depth of response that a patient goes on to achieve.

Our data support monocytoid cells (not DCs) as a primary interface relaying the PC death signal to an adaptive immune response. The greatest expression of ICD receptors LRP1 and TLR2 was seen in monocytoid cells; responders tended to have higher monocytoid cell numbers at baseline, and chemokines CCL3 and CCL4 (critical for monocytoid homing and activation) were elevated in the serum of deep responders. Monocytoid homing and activity after belantamab mafodotin treatment has also been demonstrated preclinically.^4^ These findings suggest an important role for the monocytoid population in mediating the downstream immune cascade. In solid tumors, ICD-triggered signalling typically occurs via DCs in tumor-draining lymph nodes^41^; in contrast, in MM the BM has no draining lymph nodes, which may necessitate the transmission of these signals through alternative localized abundant monocytoid populations.

Remodeling of a polymorphonuclear myeloid-derived suppressor cell (PMN-MDSC) compartment might be a possible mechansim of action of therapies affecting the TME. We saw a CD16- and PDL1-double positive cell population over-represented in responders versus non-responders at baseline, which receded with successful treatment. These cells expressed CD66b and CD11b, and segregated with low-density BMMCs on a Ficoll gradient, all characteristic of the PMN-MDSC phenotype, previously shown to be negatively prognostic in MM.^42^ The greater dominance of these cells in responders vs non-responders prior to therapy implies that belantamab mafodotin works best in patients with an immunosuppressed TME, which are those who might have the most to gain from de-repression.

T-lymphoid compartment restructuring is another putative effector mechanism. Soluble GZMB and CXCL9 were released within 24 hours after belantamab mafodotin dosing and were linked to deep responses. Preclinical investigations indicated that GZMB may be released by MM cells undergoing belantamab mafodotin-induced inflammatory cell death, but since GZMB is a canonical marker of T-and NK-cell cytotoxicity, serum GZMB could also be derived from cytotoxic T-effector cells or NK cells that are activated through the inflammatory and immunogenic nature of MM cell death.

Consistent with this, we observed remodeling of the T-cell compartment and the emergence of a CD4 GZMB-positive population in responders, a cell type associated with a good prognosis in MM and other cancers.^43, 44^ These cells were less frequent in relapsing patients, suggesting they may hold important anti-MM potential. Given the lytic nature of PC death, the fact that these cells are MHC-II-restricted cytotoxic cells is particularly relevant. Tumor antigens from dying PCs subsequently presented by antigen-presenting cells, such as monocytes/macrophages, could enable the cytotoxic activity of CD4 T-cells against PCs (which express MHC-II).

In conclusion, patients treated with belantamab mafodotin exhibited an acute inflammatory phenotype after the first dose, which our in vitro data suggest is driven by inflammatory and immunogenic cell death. This led to release of inflammatory mediators detectable in patient serum, where CCL3, CCL4, CXCL9, and GZMB may act as early biomarkers of response. We propose that the recruitment of a monocytoid-dominated response to this inflammatory background leads to T-cell restructuring, emergence of a CD4+ GZMB+ population, and loss of inhibitory cell types. The ability of the TME to remodel may be a critical determinant of clinical response to belantamab mafodotin. Following remodeling, the TME might be capable of controlling the growth of MM, leading to durable responses, even during dose holds.

### Ethics approval and consent to participate

Studies were conducted in accordance with the Declaration of Helsinki and Good Clinical Practice guidelines, and were approved by the relevant national, regional, or independent ethics committee or institutional review boards. All patients/donors provided written informed consent for use of their samples. Ethics committees overseeing the Oxford Radcliffe Biobank, HaemBio, and Oxford Musculoskeletal Biobank, all at the University of Oxford; the UCL Cancer Institute Biobank, at University College London; and the ProMMise trial, approved sample collection/use. Full details are in the Supplemental Methods.

## Data Availability

Information about GSKs data sharing commitments can be found at https://www.gsk-studyregister.com/en/.
Additional data may be available from the authors upon reasonable request. For original CyTOF data, please contact; ONT long-read scRNA sequencing data are available at Gene Expression Omnibus (GEO) under accession number GSE307660, Xenium data are available at GEO under accession number GSE307661.

## Acknowledgements

EW was supported by Cancer Research UK (CRUK grant number SEBCATP-2022/100011). Research in the UO laboratory was supported through funding from GSK (UO, KR), Innovate UK, the National Institute for Health Research Oxford Biomedical Research Centre, Cancer Research UK, the Leducq Epigenetics of Atherosclerosis Network (LEAN) program grant from the Leducq Foundation, and the Myeloma Single Cell Consortium. SGo is supported by Cancer Research UK (CRUK grant number RCCCSF-Nov21\100004). Samples were obtained through HaemBio, the Oxford Radcliffe Biobank and the ProMMise trial team, and we acknowledge expert sample preparation support from Emma Lyon, Mirian Angulo Salazar, Batchimeg Usukhbayar, Vicki Gamble, Renuka Teague, and David Maldonado-Perez. We would like to acknowledge the advice of Tony Ng who provided the initial suggestion to explore inflammatory cell death as a potential mechanism of action and for reviewing the mechanistic aspects of the manuscript. We are grateful to Taryn Mockus-Daehn for support with flow cytometry panel design and analysis, Aurelie Bornot for supporting data management and Lydia Kondyliou for sample management. Joanna Opalinska and Pralay Mukhopadhyay provided clinical development expertise to enable the DREAMM-5 and DREAMM-14 trials. Thank you to all of the patients who took part in the studies and provided permission to collect and use the samples that made these discoveries possible. Editorial support (in the form of writing assistance, including collating and incorporating authors’ comments for each draft, assembling tables and figures, grammatical editing and referencing) was provided by Elisabeth Walsby, PhD, CMPP, and Alexus Rivas-John, PharmD, at Fishawack Indicia Ltd, part of Avalere Health, and was funded by GSK.

## Author contributions

ECRW, UO, SGr were involved in the conception and design of the study, the acquisition of data and the data analysis and interpretation. DEL, SGo, AK, PM, and KR were involved in the design of the study. HAS, MHe, JW, CYW, MHt, LL, VN, WB, BM, RD, PRB, YM, DM, QH, ACA, LL, HCE, EC, YS, JH, FI, GN, and HM were involved in the acquisition of data and data analysis and interpretation. All authors contributed to the writing and reviewing of the manuscript, and have given final approval to the version to be published.

## Data availability

Information about GSK’s data sharing commitments can be found at https://www.gsk-studyregister.com/en/.

Additional data may be available from the authors upon reasonable request. For original CyTOF data, please contact; ONT long-read scRNA sequencing data are available at Gene Expression Omnibus (GEO) under accession number GSE307660, Xenium data are available at GEO under accession number GSE307661.

