## Supplemental Material for "Belantamab Mafodotin Triggers Immune Invigoration in Multiple Myeloma Via Inflammatory and Immunogenic Cell Death"

**Supplemental Methods**

**Treatment of multiple myeloma (MM) cells:** NCI-H929 (ATCC), JJN3 (GCMCC), RPMI-8226 (ATCC) or BMMCs from MM patients (obtained from Discovery Life Sciences) were seeded into 96-well plates at a density of 1 x 10^5^ (HMCL) or 2 x 10^5^ (BMMC) cells/well. HMCLs were seeded in RPMI 1640 Medium (ATCC modification; Gibco) for NCI-H929 and RPMI-8226, and DMEM/IMEM (1:1 ratio, Gibco) for JJN-3,supplemented with 10% (H929 and RPMI-8226) or 20% (JJN-3) fetal bovine serum (FBS; Gibco), 0.05 mM 2-mercaptoethanol (Gibco; NCI-H929 only), and 1% penicillin-streptomycin (Gibco). Unsorted BMMCs seeded in RPMI 1640 with GlutaMax (Gibco) supplemented with 10% fetal bovine serum (FBS; Gibco), and penicillin-streptomycin (Gibco). Cells were incubated for 16 (BMMC) 48- or 72-hours (HMCL) with varying concentrations of belantamab mafodotin (GSK), monomethyl auristatin F (MMAF) (Selleck Chem) or negative controls, daratumumab (Selleck Chem), IgG1 (Selleck Chem), and DMSO (Sigma-Aldrich). BMMCs were screened by flow cytometry for the presence of BCMA+ cells and only samples with viable BCMA+ cells after 16-hour incubation were included. Post-incubation, plates were centrifuged at 300 *g* for 10 minutes at 4°C. Supernatants were collected and stored for subsequent analysis, while the cell pellets were either processed for RNA extraction or prepared for flow cytometry analysis.

**Analysis of cell death markers by flow cytometry:** Cell pellets were washed with 150 μL PBS and resuspended in 100 μL of a live/dead staining mix (Zombie Aqua, Biolegend), diluted at a ratio of 1:400, combined with an FcR Block solution (Miltenyi) diluted at 1:100. Samples were incubated for 20 minutes at 4°C, protected from light. Plates were centrifuged at 500 × g for 3 minutes at 4°C, and the supernatant discarded. 50 μL of primary antibody against surface antigen or fluorescence-minus-one (FMO) controls (**Supplemental Table 1**) were incubated with cells for 60 minutes at 4°C, then washed twice with 100 μL of cell staining buffer (CSB, Biolegend), resuspended in 100 μL of fixation/permeabilization buffer (eBioscience) and incubated for 20 minutes at room temperature (RT). Post-incubation, plates were centrifuged at 500 × g for 3 minutes at 4°C, washed once in CSB, followed by 1 wash in permeabilization buffer and resuspended in 50 μL of antibodies specific for intracellular antigens diluted in intracellular staining/permeabilization buffer (eBioscience). After 60 minutes incubation at 4°C, cells were washed once with 100 μL of permeabilization buffer and once with 100 μL CSB. For secondary staining, cells were incubated with 50 μL of secondary antibody staining solution (**Supplemental Table 1**) for 40 minutes at 4°C, protected from light. Finally, cells were washed twice with 100 μL of CSB buffer, resuspended in 150 μL of CSB and data were acquired on the Cytek Aurora cytometer. Data were analyzed using FlowJo 10.10 software (Flow Jo, LLC). For the analysis of plasma cells (PC) from MM patients, we used a sequential gating strategy where we first identified live, CD3-, CD14-, CD16-, CD11c- population and the CD40+CD45- cells contained within it as PC and gated on BCMA+ cells to identify BCMA+ PC. Statistical analysis was carried out in R version 4.4.1. For each marker, a binomial proportions model was used for data analysis. For NCI-H929 cells, the models contained a fixed effect for treatment group and response was the proportion of total live cells that are also positive for the specified marker. For primary cells, the models contained both a fixed effect for treatment group and a random effect for donor, and response was the proportion of BCMA+ cells that are also positive for the specified marker. Mean responses and standard error of the means for each group were computed, and linear contrasts were derived which provide odds ratios comparing each treatment group to its respective control. For each marker, the Benjamini-Hochberg multiplicity correction was applied to control the FDR set at 5%.

**Flow cytometry analysis of peripheral blood samples:** DREAMM-5 is a phase 1/2, randomized, open-label, platform study designed to evaluate the effects of belantamab mafodotin in combination with other anti-cancer drugs in patients with relapsed/refractory MM. Peripheral blood mononuclear cells (PBMC) samples from baseline, Cycle 2 Day 1, or Week 5 (C2D1+Week5) and End of Treatment (EoT) were available for 33 patients from DREAMM-5 sub-studies 3 (belantamab mafodotin with nirogacestat), 6 (belantamab mafodotin, nirogacestat, lenalidomide, and dexamethasone), and 7 (belantamab mafodotin, nirogacestat, pomalidomide, and dexamethasone). Whole blood samples were drawn in Sodium Heparin Vacutainer collection tubes (Beckton Dickinson, Franklin Lakes, NJ). Antibodies (**Supplemental Table 2**) were added to the whole blood sample, incubated, lysed, and washed per manufacturer guidelines, and data were collected using BD FACSCanto II (BD Biosciences, Franklin Lakes, NJ). Cellular phenotypes were defined as outlined in **Supplemental Figure 9**. Replicate subject measurements within each time point were summarized using the median value. Statistical analysis was performed using the Linear Mixed Model (LMM) to model the longitudinal data (lmer) function from the lme4 package in R). In the LMM all data were transformed using Yeo-Johnson transformation and the p-values of the comparisons are adjusted by Tukey-Kramer method within each marker of interest.

**Bulk RNA sequence analysis of cell line transcriptomes:** NCI-H929 cells (1 x 10^5^ per well of a 96 well plate) were treated at 37°C with 1 or 10 µg/mL belantamab mafodotin, 5 or 10 µg/mL MMAF, 5 µg/mL IgG1, or DMSO for 48 hours, with 4 replicates per condition. RNA was extracted using a KingFisher Apex system according to manufacturer’s protocol. QC was performed using TapeStation before samples were further processed (Azenta Life Sciences) to conduct strand specific bulk RNAseq (poly-A selection, Illumina 2x150 bp paired-end sequencing, 100 million reads). Sequencing reads were soft trimmed using trimmomatic, then aligned to GRCh38 using STAR mapper, then de-duplicated using Picard MarkDuplicates and gene expression for genes from Ensembl v96 detected using Subread featureCounts. Principal component analysis showed that the first two Principal Components (PC) explained 92% of variability without signs of experimental batch effects. Differential gene expression analysis was performed using DESeq2.^1^ Contrasts were extracted using the appropriate control (DMSO for MMAF and IgG1 for belantamab mafodotin). Log2 fold changes were moderated using Ashr.^2^ Multiple testing correction was performed by controlling the FDR using Independent Hypothesis Weighting.^3^

The detected log-fold changes were categorized into 6 positive integer buckets capped at 5, 6 negative integer buckets capped at -5 and 3 additional categories labeled ‘n.s.’ (gene expressed but not significant with FDR >0.05*).* For the pathway analysis, count libraries were first normalized using the TMM method (trimmed mean of M-values), then analysed with voom before the method fry was applied (edgeR v4, Chen et al., NAR, 2025). Gene sets were obtained from Reactome DB stored in the 'Molecular Signatures Database' (MSigDB) obtained by the R package msigdbr. Necroptosis used Reactome ID: R-HSA-5213460; n=34 genes and Pyroptosis Reactome ID: R-HSA-5620971; n=30 genes*.*

**Analysis of secreted inflammatory factors:** Supernatant or patient serum was evaluated using a U-plex custom 10 plex kit (Mesoscale Discovery, MSD #K15235N-2) to quantify secreted ICD-associated inflammatory mediators (IL8, Granzyme B, IL16, CCL3, CCL4, IL18, IL1β, CXCL9, CCL2 and CXCL1) or HMGB1 ELISA (Cloud Clone Corp #SEA399Hu). All assays were run following the manufacturer’s supplied protocols. Concentrations were calculated using a standard curve (ran in duplicate) for each analyte using MSD software (Discovery workbench version 4.0.13) or R for HMGB1 ELISA. Concentration values were corrected for the dilution factor, and only values within the linear range of the standard curve were included in the analysis; values above the upper limit of detection (ULOD) or below the lower limit of detection (LOD) were clearly indicated. Statistical analysis was carried out in R version 4.4.1. For each of the secreted inflammatory mediators from NCI-H929 or BMMCs, a Bayesian Tobit model was implemented. For NCI-H929 cells, the models contained a fixed effect for treatment group, and for primary cells, the models contained both a fixed effect for treatment group and a random effect for donor. For NCI-H929 cells, the Benjamini-Hochberg multiplicity correction was applied to control the FDR at 5%. For analysis of patient serum, LMM and Bayesian Tobit models (for markers with values either exceeding the ULOD or below the LLOD) were used to assess whether the longitudinal profiles of inflammatory markers changed over time and whether these profiles differed between response groups. Pairwise comparison p-values from the LMM were adjusted with the Tukey-Kramer method, while interaction p-values between response groups and time points were adjusted for multiple testing of 9 markers using the Benjamini-Hochberg FDR method.

**Proteomic analysis:** Cell culture supernatants were analyzed using four Olink Explore panels: Explore 384 Inflammation, Inflammation II, Oncology, and Neurology. The analysis was performed according to manufacturer’s instructions (Olink protocol v4.0, 2024-04-16). Final library pools of 2 panels each were combined on a SP flowcell 100 cycles kit (Illumina). Libraries were sequenced on a NovaSeq 6000 instrument (Illumina), read mode 24-0-0-0 using a custom recipe provided by Olink. Raw data was processed with ngs2counts software (v.4.7.1) to generate counts per analyte per sample. The NPX Explore software (v.3.9.0) was used for quality control and normalization. Olink protein levels are expressed in Normalized Protein eXpression (NPX) units (log2 scale), with a 1 NPX difference translating into a doubling of protein concentration. The lower limit of detection (LOD) as provided by the NPX Explore software was utilized for censoring: proteins were retained for analysis if NPX values for more than 50% of all replicates within one group were above LOD. Statistical analyses were conducted using R version 4.3.1, with linear models fitted separately to each protein using the lm() function. The emmeans() function in the emmeans package was used to perform post-hoc multiple pairwise comparisons of mean abundance among groups. P-values were adjusted for multiple testing using the Benjamini-Hochberg FDR method

**Multiplex immunofluorescence imaging:** Paired formalin fixed paraffin embedded (FFPE) biopsies were collected at screening and on cycle 2 day 15 or cycle 3 day 1, respectively from 6 patients from DREAMM-5 sub-study 5 (belantamab mafodotin + isatuximab) and 4 patients from substudy 2 (belantamab mafodotin + feladilimab). FFPE blocks were surface-decalcified for at least 30 minutes on ice with EDTA decalcifier (PRC), sectioned to 4 µm using a Sakura Tissue-Tek AutoSection and mounted on SuperFrost Ultra Plus™ GOLD Adhesion Slides (Epredia). Slides dried overnight at 37°C were stored at RT. Samples were baked, dewaxed, and treated with BOND Epitope Retrieval Solution 2 (Leica) using Leica BOND RX auto stainer. Imaging was performed using the Leica Cell DIVE system through 7 rounds of fluorescent-label antibody staining (**Supplemental Table 3**), imaging, and dye inactivation with 3% hydrogen peroxide (Merck) and 0.1 M sodium bicarbonate (Merck). Primary antibody stained for 1 hour at 37^o^C with 10-minute RT DAPI stain (Thermo) with Leica BOND RX auto stainer. Leica Cell DIVE automated image alignment used DAPI signal. Unconjugated primary antibodies were conjugated using the FlexAble 2.0 CoraLite® Plus 555 Rabbit IgG labeling kit (Proteintech). Image analysis was performed using custom Python scripting. For each sample, individual channel images for CCR7, CD45RA, and CD4 were corrected for an observed vignetting artefact using the BaSiC package.^4^ Nuclei were segmented from the DAPI channel with CellPose^5^ features were extracted from the image stack using the 3px perimeter dilation of obtained nuclear masks. Cells were typed for marker positivity through manual intensity thresholding per channel and passed through a hierarchical gating strategy to assign cell type. Cell type density was calculated by Otsu thresholding the DAPI channel of each image to determine tissue area using the marker combination described in **Supplemental Table 4**. To ensure purity of the analyzed MM/PC fraction and negate off-target binding of each individual antibody, MM/PC were classified as CD138+ BCMA+ double positive. Cells single-positive for CD138 or BCMA were excluded from analysis. Generalized least squares models were used to assess the percent cell type, density of cell type or biomarker intensity change between screening and on-treatment time points, and the interaction effect of time and response on the percent cell type or marker expression changes. Log-transformed percent of cell type, log-transformed cell density or log-transformed median intensity was used in the analysis.

**Bone marrow samples from belantamab mafodotin-treated patients:** Bone marrow aspirates from patients treated with belantamab mafodotin monotherapy were acquired either: as part of routine care and stored within one of the Oxford Radcliffe Biobank (REC reference 19/SC/0173), HaemBio (REC reference 24/EM/0060), or UCL Cancer Institute Biobank (REC reference 20/YH/0088); or, within the ProMMise trial (REC reference 21/WS/0065) with patient consent for future research. Whole bone marrow aspirate was passed through a CD138 bead selection kit (EasySep from StemCell); the flow-through (CD138- fraction) was separated on a Ficoll gradient, and the buffy coat was isolated, counted, and cells were viably frozen in DMSO/foetal calf serum (10/90, v/v); the cells retained on the beads were eluted, counted and ~ 500,000 were pelleted in buffer RLT (Qiagen) viably frozen and stored in liquid nitrogen until use. FFPE blocks were retrieved through the Oxford Radcliffe Biobank (REC reference 19/SC/0173). Vials with BM aspirates were defrosted in small batches in a 37°C water bath for approximately 1 minute, then placed on ice and transferred drop-wise into 4^o^C RPMI 1640 medium supplemented with 1% glutamine (Gibco) (concentration), and 10% FCS (v/v). Samples were washed twice by centrifugation at 300 × g for 5 minutes. Cells were resuspended in sterile PBS up to a concentration of 1 million per mL and passed through 70 µm filter tips (Bel-Art Flowmi). Samples were immediately proceeded to parallel mass cytometry staining and single-cell transcriptomics workflows.

**Bone marrow samples from non-malignant controls:** Femoral head and necks acquired during elective arthroplasty for non-malignant conditions were taken directly for processing, where trabecular bone marrow was washed with ice cold PBS and agitated. The resulting eluate was spun on a Ficoll gradient and the buffy coat taken forward for long read scRNA sequencing. All samples were obtained from the Oxford Musculoskeletal Biobank (OMB), University of Oxford, with informed donor consent and ethical approval (REC reference 24/SC/0224).

**Mass cytometry:** Up to 3 million cells per sample were transferred to a 5 mL round-bottomed polystyrene tube, and washed twice in Maxpar Cell Staining Buffer (CSB; Standard BioTools) with 5 minutes centrifugation at 300 × g. Prior to intracellular permeabilization, cells underwent an additional 1.6% formaldehyde fixation. Rhodium live/dead staining was included in the surface antibody staining reaction. Normalized .fcs files were uploaded into Cytobank flow cytometry software and processed as previously described.^6^ Singlet events were exported as .fcs files and processed in R (version 4.4.1, R Core Team 2024). QC and visualization was performed with CATALYST; ^7^ batch correction with CyCorrect and differential abundance analysis with MiloR.^8^

***Supplemental Figure 1. Patients maintain continued clinical benefit during prolonged belantamab mafodotin dose holds***
Patients treated with belantamab mafodotin monotherapy under the named patient program (NPP) at a single institution had therapy administration data and paraprotein or light chain results recorded. In some cases, especially very good partial response or deeper partial response, the burden of heavily pre-treated disease would remain at stable levels despite 3 months or more off treatment (licensed inter-dose interval is 3 weeks). Each panel represents a single patient. Top figure per panel is overall therapy timeline, where black points mark a biopsy timepoint. Lower figure is an expanded view of the belantamab mafodotin monotherapy therapy window, where: blue line represents disease burden according to primary y-axis (paraprotein or difference between involved and uninvolved light chain); purple dot represents dose of belantamab mafodotin administered, according to secondary y-axis; black vertical bar corresponds with bone marrow biopsy. Note different scales per patient. Belamaf, belantamab mafodotin.

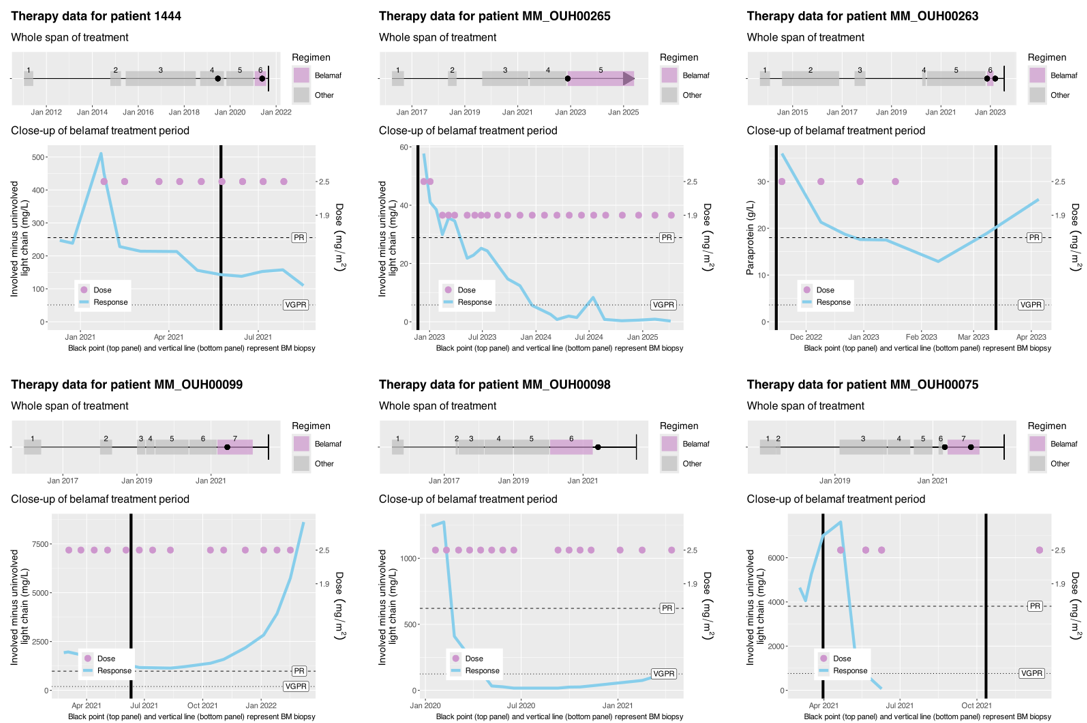

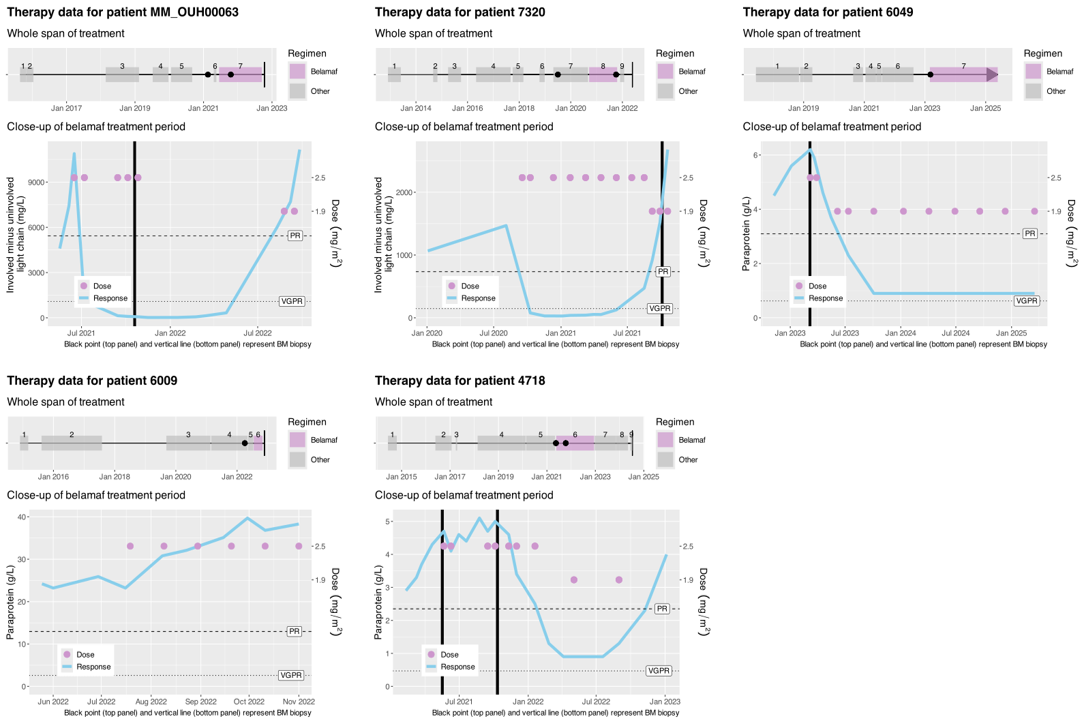

***Supplemental Figure 2. Belantamab mafodotin and MMAF free payload trigger induction of pyroptosis and necroptosis pathways***

(A) ICD and inflammatory cell death markers measured by flow cytometry in JJN3 and RPMI-8226 after 48 hours’ treatment with either belantamab mafodotin or MMAF. (B) MA plot of RNAseq results of MMAF-treated NCI-H929 (either 5 µg/mL or 10 µg/mL for 48 hours vs DMSO control), showing mean log2 expression on x-axis and moderated log2 fold change on y-axis. Genes meeting the significance threshold (FDR < 0.1) are shown in green. Genes implicated in cell death pathways are highlighted in red for necroptosis (right) and pyroptosis (left). (C) Primary patient BMMCs were incubated with MMAF for 16 hours and cell surface markers assessed by spectral flow. For each marker, a binomial proportions model was used to compare treatment response vs IgG1 vehicle control and the BH multiplicity correction was applied to control the FDR set at 5%. Graph represents amalgamated data from all 7 donors where the colors represent donors and the dots represent replicates. Error bars are standard error of the mean, ***P<0.001; **P<0.01; *P<0.05. (D) Heatmap comparing the Log2 odds ratio changes in cell surface marker by flow cytometry following treatment of 7 different MM donor BMMCs with belantamab mafodotin, MMAF, or daratumumab negative control. White (n.s) = not significant. Belamaf, belantamab mafodotin.

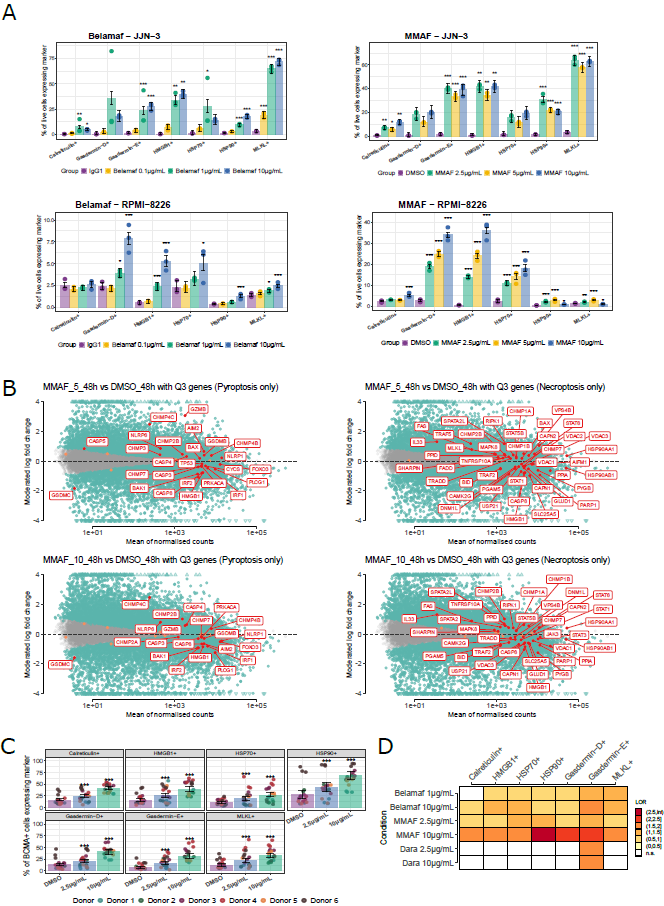
***Supplemental Figure 3. MMAF free payload triggers chemokine release***
(A) Volcano plots visualizing the Log2 fold changes in analyte concentrations in the supernatant from NCI-H929 cells treated for 48 and 72 hours with free-MMAF at 2.5, 5, and 10µM. Proteins showing a Log2 fold change of at least 1 and an adjusted P-value of <0.05 are considered significant and are colored in red. (B) MA plot of RNAseq results for *CCL3*, *CCL4*, *GZMB*, and *CXCL8* of either belantamab mafotodin or MMAF-treated NCI-H929 cells (10 µg/mL for 48 hours vs either IgG or DMSO control), showing mean log2 expression on x-axis and moderated log2 fold change on y-axis. Genes meeting the significance threshold (FDR < 0.1) are shown in green. (C) Concentration of selected pro-inflammatory molecules in the supernatant of NCI-H929 cells treated with MMAF as measured by MSD. Dotted line indicates lower limit of detection of the assay in the linear range. Error bars are standard error of the mean. For each of the secreted inflammatory mediators, a Bayesian Tobit model was implemented to compare treatment vs IgG control ***P<0.001; **P<0.01; *P<0.05. Belamaf, belantamab mafodotin.

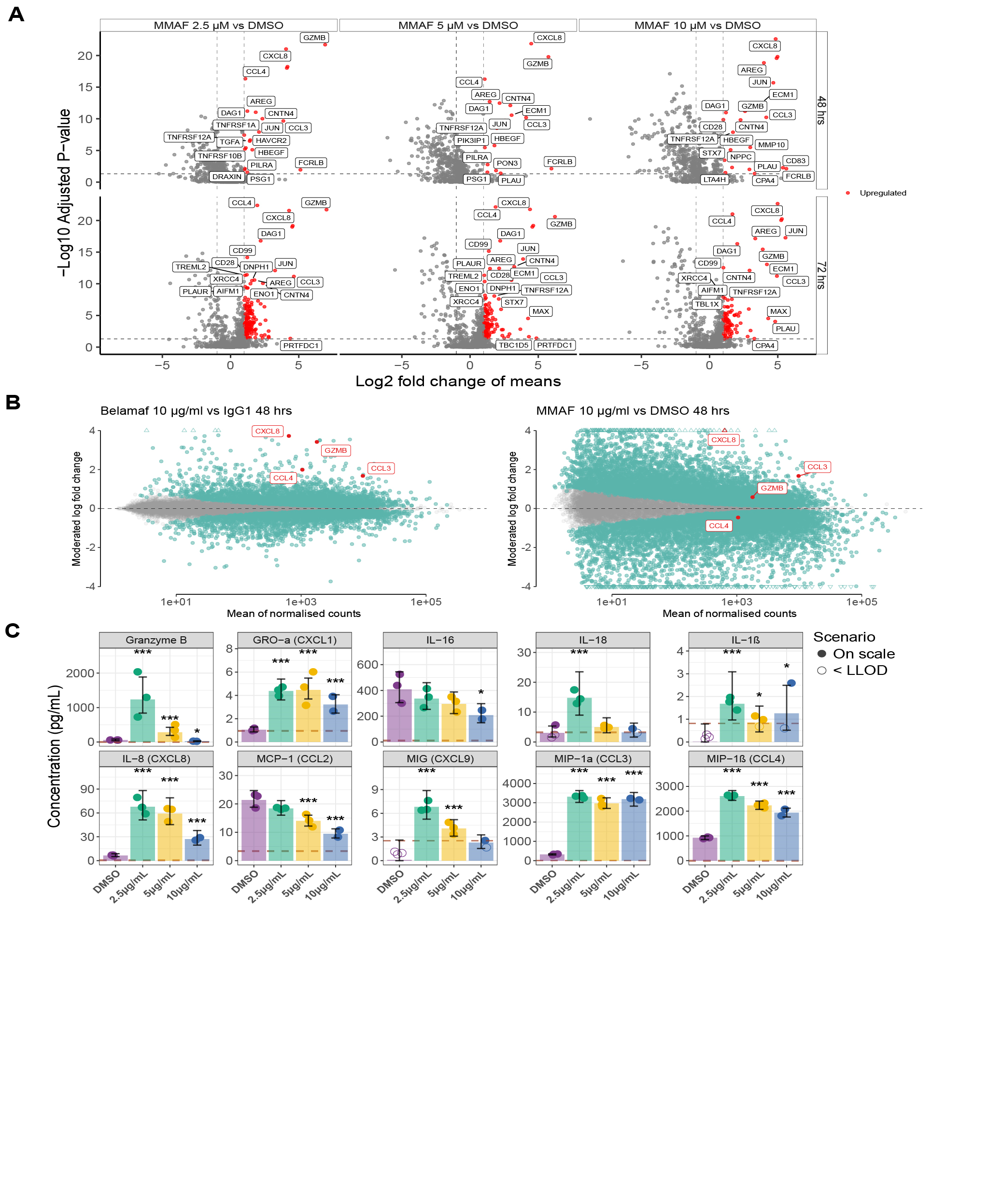

***Supplemental Figure 4. Spatial transcriptomic analysis of bone marrow trephines***
Samples from patients in the named patient program who received belantamab mafodotin monotherapy: (A) Xenium images of MM075 pretreatment (left panel) and follow-up (right panel) with the selected tiles (500 μm x 500 μm) applied in the study. The representative spatial tiles were marked in red. (B) Images of representative spatial tile from patient MM075 at pretreatment (left panel) and follow-up (right panel), with colors indicating distinct cell types. (C) Average cell composition (percentage)in selected spatial tiles (n=4 per group) from patient MM075 at pretreatment (left panel) and follow-up (right panel); error bars represent 1 standard deviation from the mean. (D) Heatmap showing the activity of inferred signaling pathways in patient MM075 at pretreatment and follow-up. (E) Bar plot depicting the strength of individual interactions in the MHC-I signaling in patient MM075, comparing pretreatment (red) and follow-up (blue) samples. (F) Dot plot of outgoing (orange-red) and incoming (purple) signaling pathways between selected cell clusters in pretreatment (left) and follow-up (right) samples from MM075***.*** Dot size indicates the relative contribution of the cell cluster to the signaling pathway.

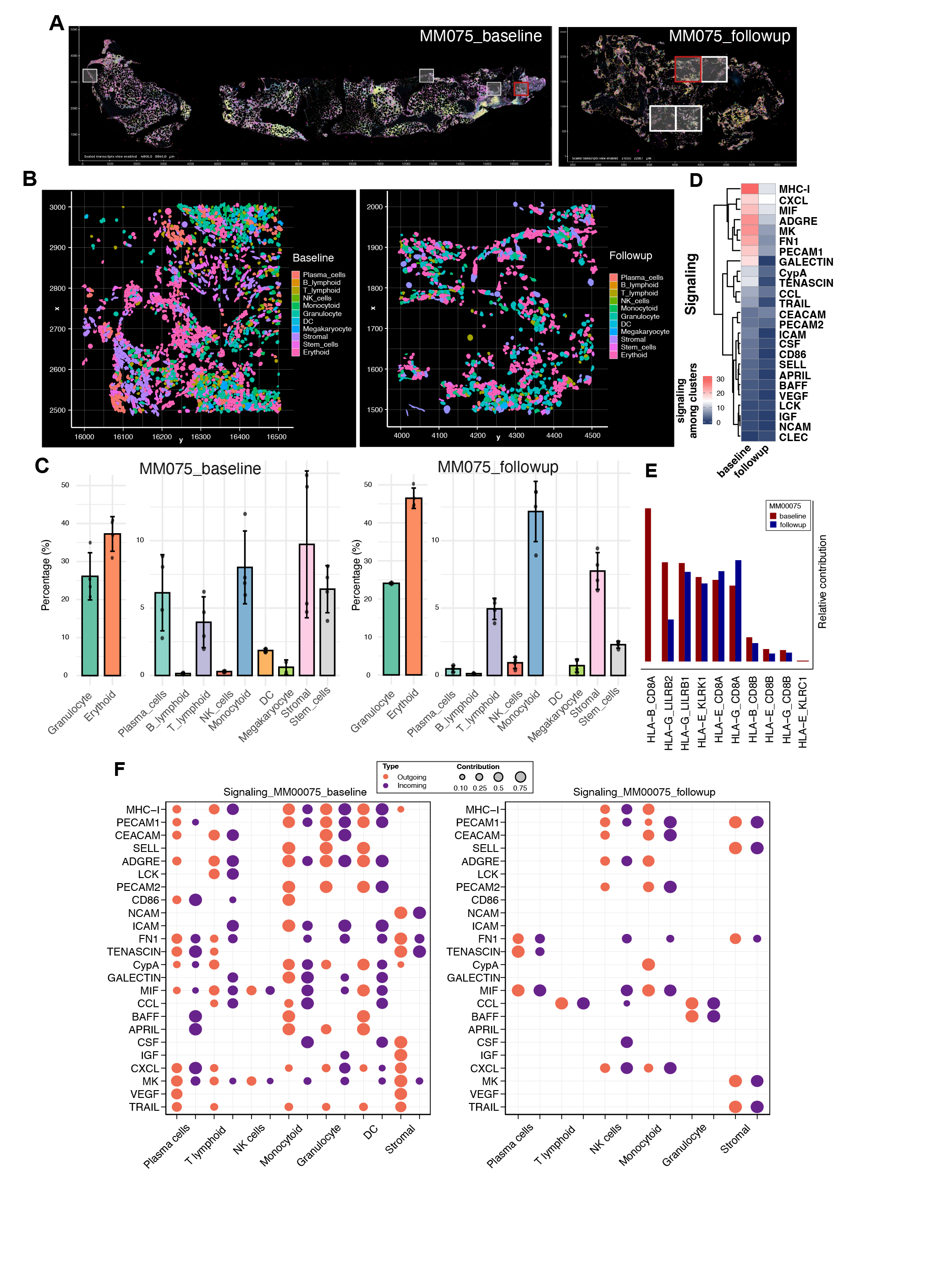

***Supplemental Figure 5. The bone marrow of responding patients shows signs of invigoration over time***
(A) Representative multiplex immunofluorescence image from trephine of a DREAMM-5 patient. (B) Relative proportions of cell types in patients between screening and follow-up, with responding patients on left and non-responding patients on the right, from multiplex immunofluorescence imaging. S=Screening timepoint, T=on-treatment. (C–E) Analysis of multiplex immunofluorescence imaging where generalized least squares was used to assess the percent cell type, density of cell type or biomarker intensity change between screening and on-treatment time points within response groups, and the interaction effect of time and response on the percent cell type or marker expression changes. (C) Relative changes in ICD and pyroptosis markers in responders vs non-responders, as determined by multiplex immunofluorescence. (D) Relative changes in cell populations in responders vs non-responders, as determined by multiplex immunofluorescence. (E) Relative changes in pro-inflammatory CD11b- and anti-inflammatory CD11b+ macrophage populations by responders.

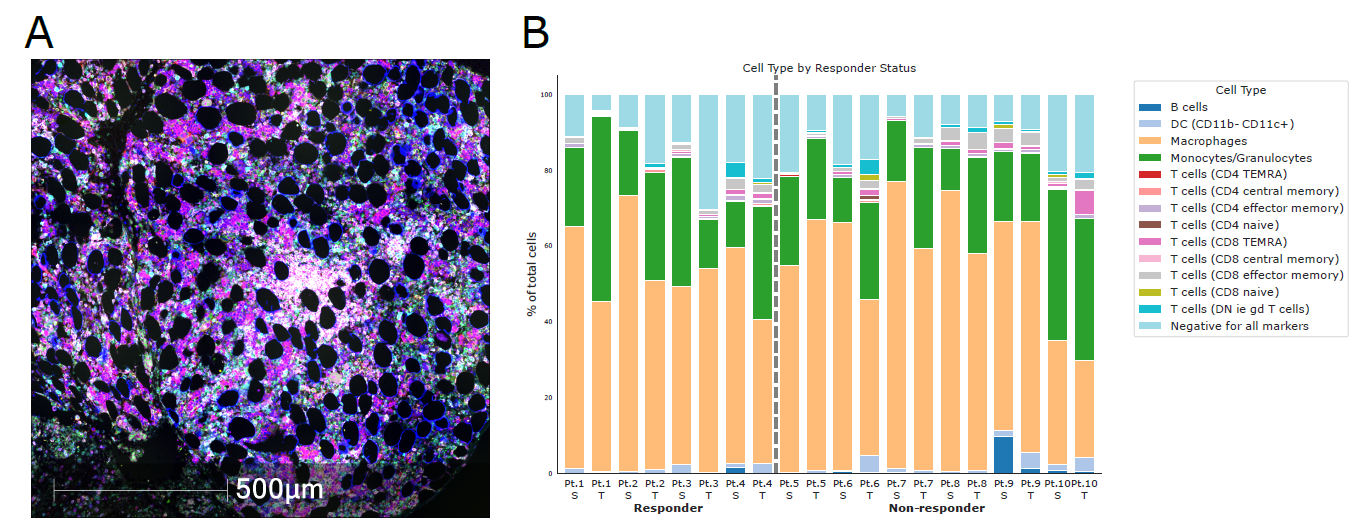

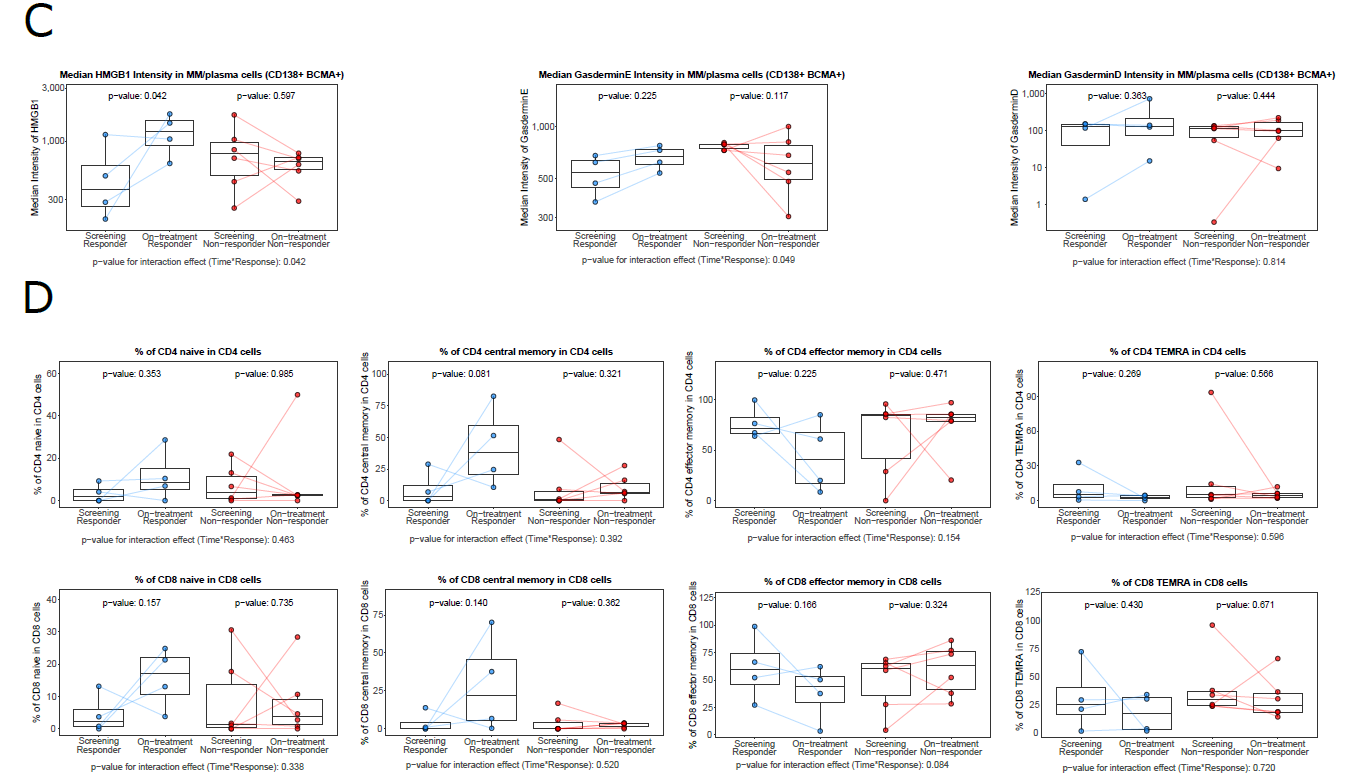

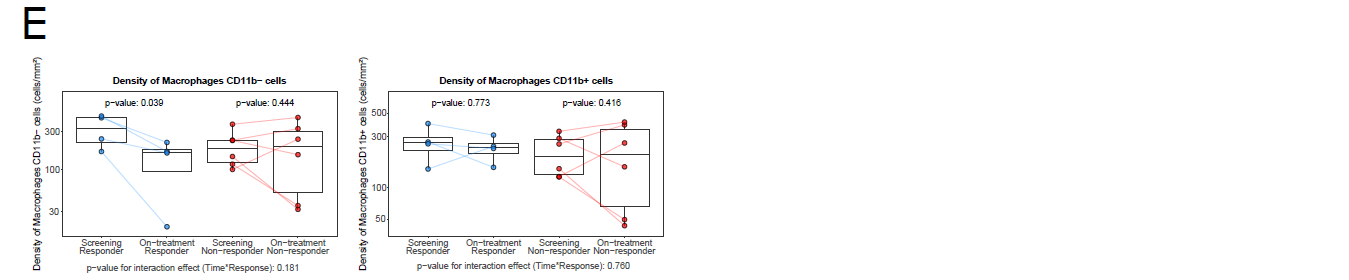

***Supplemental Figure 6. Analysis of peripheral blood indicates changes in adaptive immunity***
Belantamab mafodotin shows remodeling of the T-lymphoid landscape in peripheral blood of treated patients. Proportions of immune cells at baseline, C2D1, and EoT from 33 patients in the DREAMM-5 study. Replicate subject measurements within each time point were summarized using the median value. A linear mixed model was used to test changes over time. EoT, end of treatment and values reaching significance of p≤0.05 are highlighted in red.
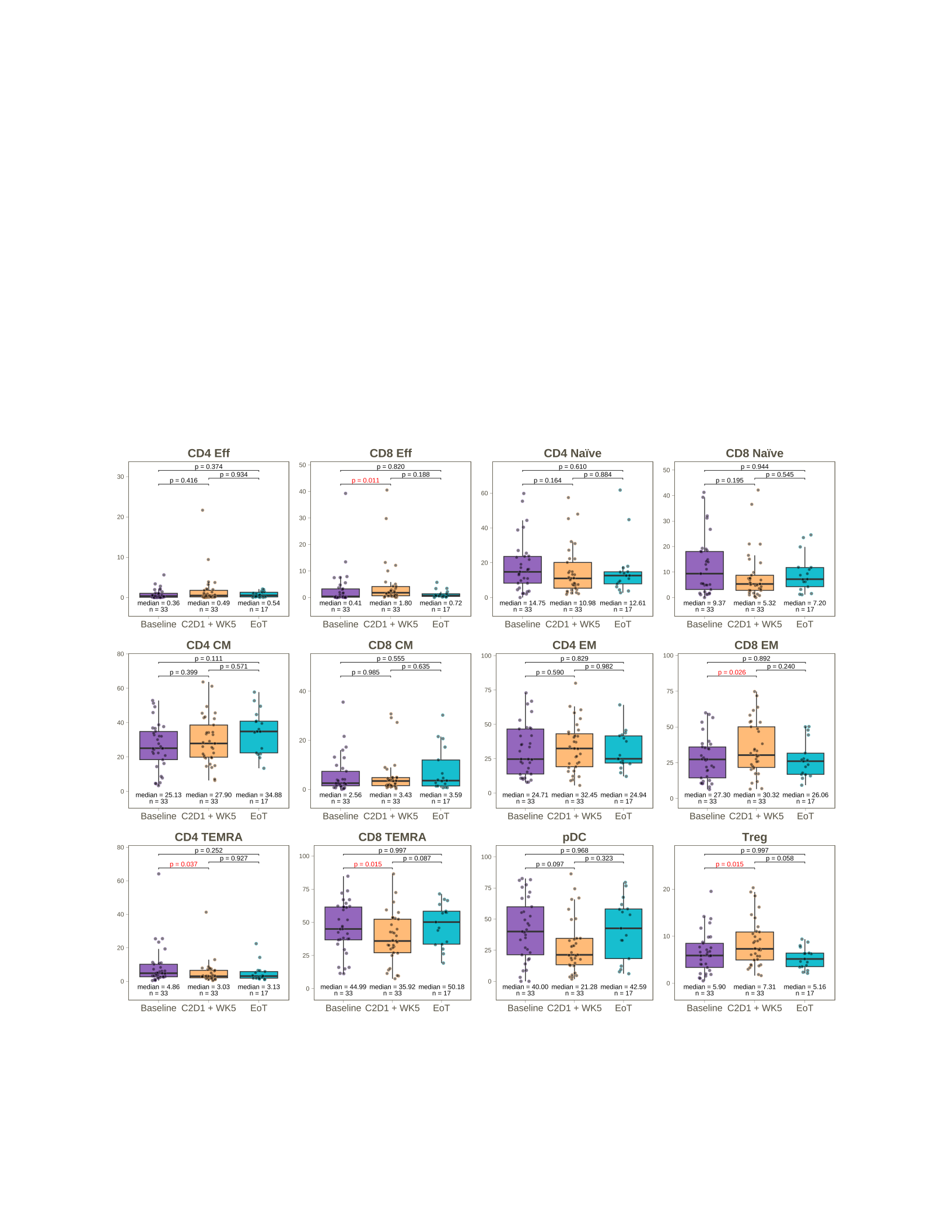

***Supplemental Figure 7. Granzyme B levels in CD4-positive, GZB-positive cells are equivalent to cytotoxic CD8 cells***

Samples from patients in the named patient program who received belantamab mafodotin monotherapy: scatter plots from normalized mass cytometry analysis of patient bone marrow samples. Arcsinh-transformed expression values for CD4 and granzyme B are shown for each population of the T-cell compartment.

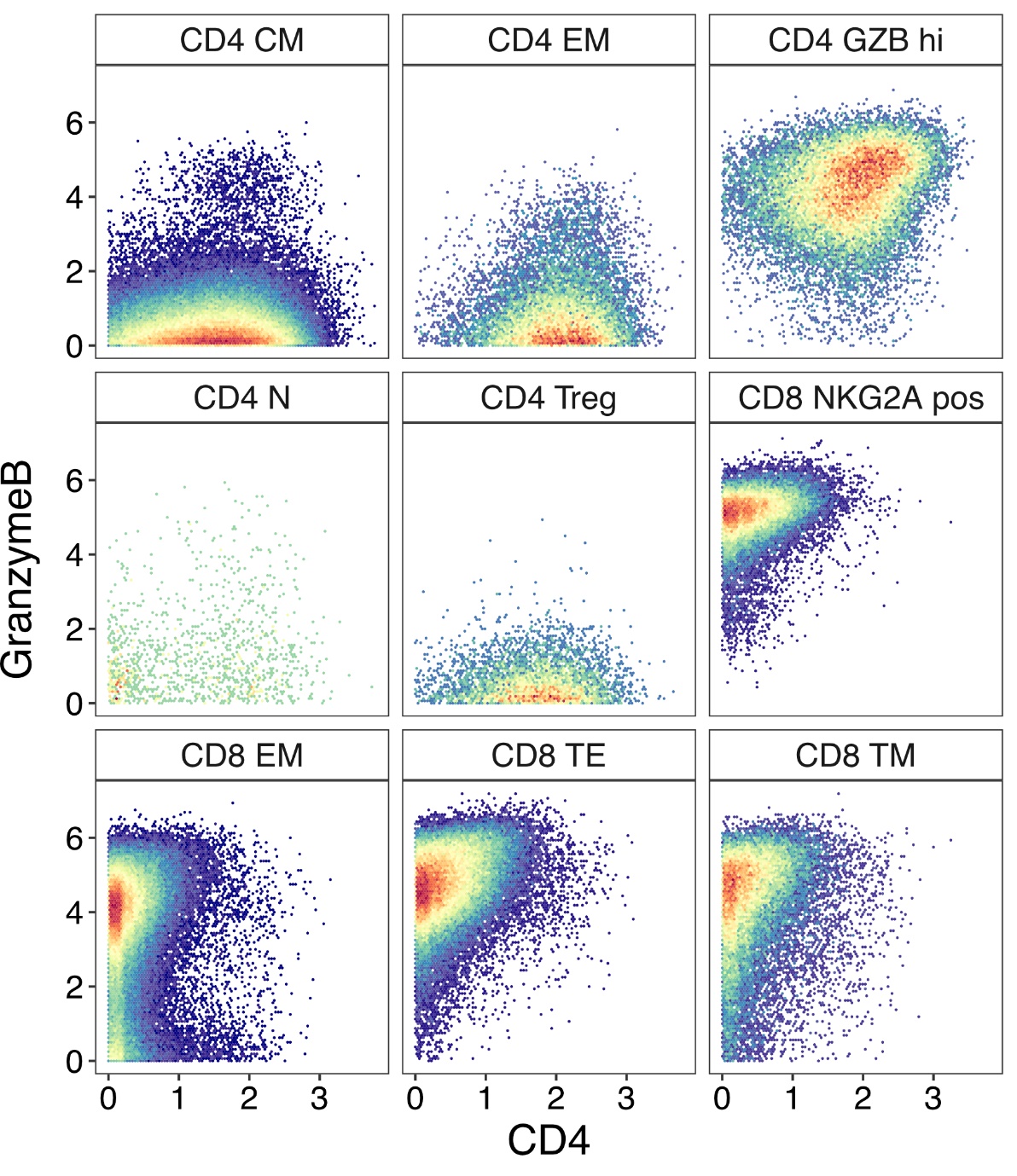

***Supplemental Figure 8. Matrix of bone marrow samples available from monotherapy-treated patients***
Samples from patients in the named patient program who received belantamab mafodotin monotherapy: long-read scRNA seq and mass cytometry were acquired in parallel for each sample. See Supplemental Figure 1 for relative timings of biopsies for responding patients. T, parallel trephine data acquired by *in situ* spatial transcriptomics.

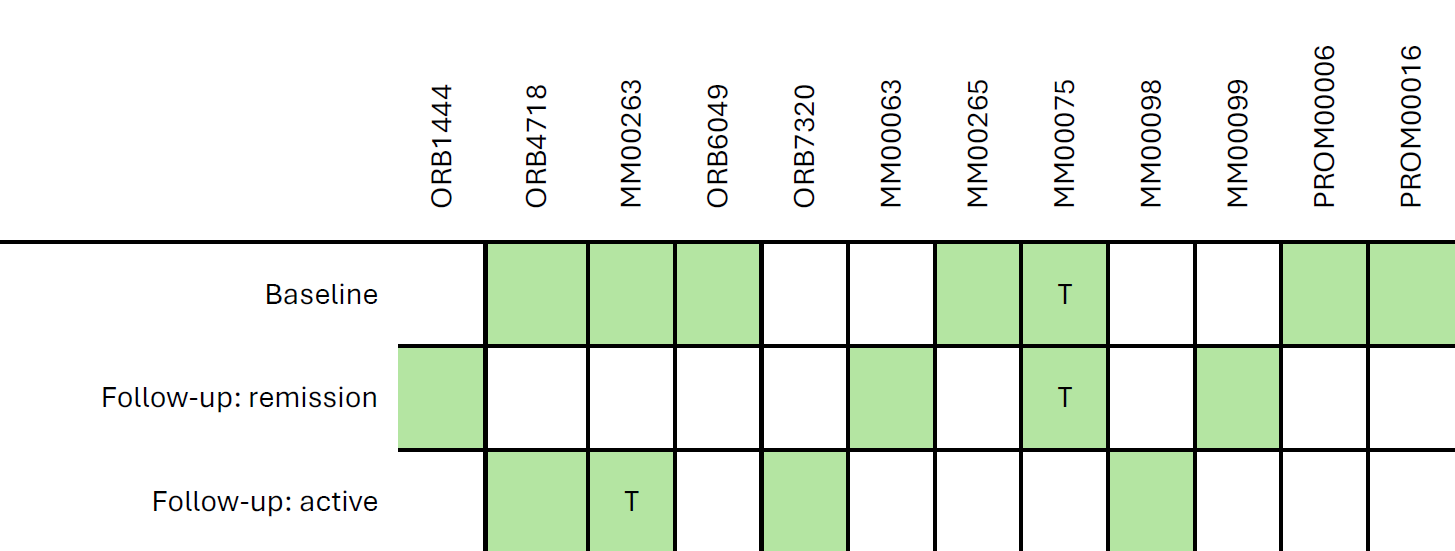

R, responder; NR, non-responder; T, parallel trephine data acquired by in situ spatial transcriptomics.

***Supplemental Figure 9. Gating strategy and cell phenotypes for flow cytometry of peripheral blood from patients in DREAMM-5***

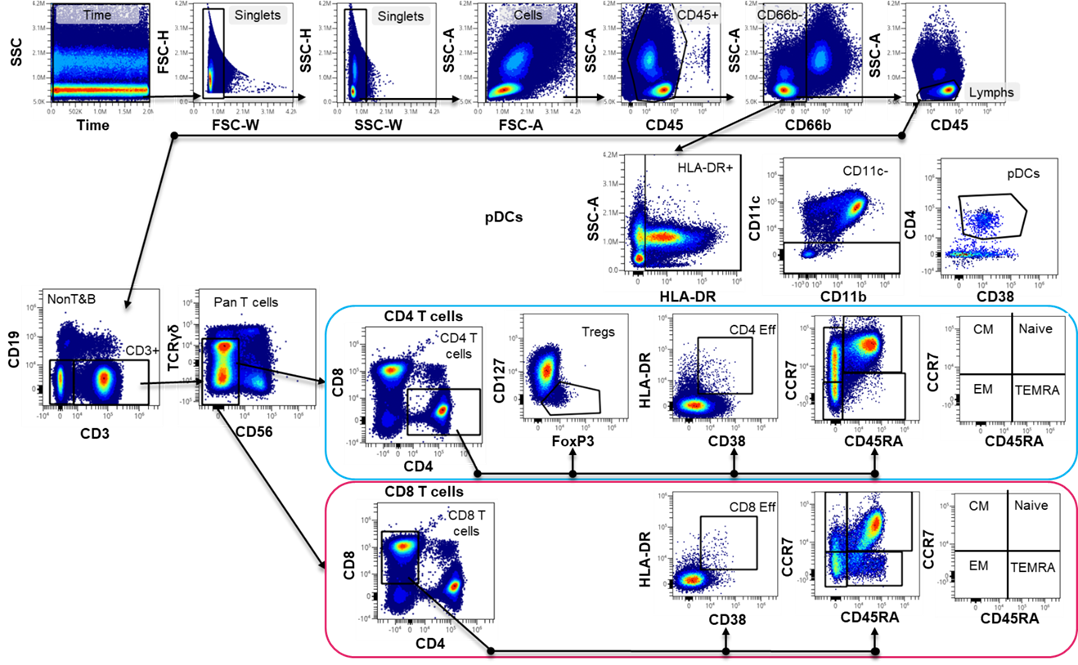

***Supplemental Table 1. Antibody marker panel used for ICD detection by spectral flow cytometry***

| **Molecule** | **Clone** | **Flouro-chrome** | **Panel Cell Line** | **Panel BMMCs** | **Source** | **Cat number** |
| --- | --- | --- | --- | --- | --- | --- |
| Calreticulin | EPR3924 | AF647 | ✓ | - | Abcam | ab196159 |
| Calreticulin | 1G6A7 | APC | - | ✓ | Novus | NBP1-47518APC |
| CD38 | HB-7 | BV650 | ✓ | - | BioLegend | 356620 |
| CD138 (Syndecan-1) | MI15 | BV785 | ✓ | ✓ | BioLegend | 356538 |
| HMGB1 | EPR3507 | AF405 | ✓ | ✓ | Abcam | ab206895 |
| CD269 (BCMA) | 19F2 | APC/Fire 750 | ✓ | ✓ | BioLegend | 357516 |
| HSP90 | EPR16621-67 | AF488 | ✓ | ✓ | Abcam | ab223467 |
| Cleaved N-terminal Gasdemin (GSDMD) - BSA and Azide free | EPR20829-408 |  | ✓ | ✓ | Abcam | ab255983 |
| DFNA5/GSDME - N-terminal | EPR19859 |  | ✓ | ✓ | Abcam | ab215191 |
| HSP70 | EPR17677 |  | ✓ | ✓ | Abcam | ab250639 |
| MLKL | 954724 | DyLight 594 | ✓ | ✓ | Novus | MAB91871DL594 |
| CD91 | A2MR-α2 | BUV615 | - | ✓ | BD Bioscience | 751452 |
| CD86 | BU63 | BUV737 | - | ✓ | BD Bioscience | 748376 |
| CD80 (B7-1) | 2D10.4 | BUV805 | - | ✓ | BD Bioscience | 751733 |
| CD11c | B-ly6 | BV480 | - | ✓ | BD Bioscience | 566135 |
| CD40 | HB14 | BV605 | - | ✓ | BD Bioscience | 569044 |
| CD16 | 3G8 | BUV661 | - | ✓ | BD Bioscience | 750284 |
| CD14 | M5E2 | BV650 | - | ✓ | BioLegend | 301836 |
| CD45 | HI30 | PerCP | - | ✓ | BioLegend | 304026 |
| CD3 | UCHT1 | BUV395 | - | ✓ | BD Bioscience | 563546 |
| CD8 | RPA-T8 | BUV563 | - | ✓ | BD Bioscience | 612914 |
| Zombie Aqua™ Fixable Viability Kit |  |  | ✓ | ✓ | Biolegend | 423102 |
| PerCP/Cy5.5® Conjugation Kit - Lightning-Link® (Gasdermin D conjugate) |  | PerCp/Cy5.5 | ✓ | ✓ | Abcam | ab102911 |
| PE/Cy7® Conjugation Kit - Lightning-Link® (HSP70 conjugate) |  | PE-Cy7 | ✓ | ✓ | Abcam | ab102903 |
| Goat Anti-Rabbit IgG H&L (PE) preadsorbed (GSDME secondary) |  | PE | ✓ | ✓ | Abcam | ab72465 |

***Supplemental Table 2. Antibody marker panel used for PBMC immune profiling by spectral flow cytometry***

| **Fluorochrome** | **Control panel** | **Full Panel** | **Clone & Cat Number** | **Source** |
| --- | --- | --- | --- | --- |
| PE | FOXP3 | FOXP3 | Clone: 259D Cat# 320208 | BioLegend |
| YG584 | CD4 | CD4 | Clone: SK3  Cat# R7-20041 | Cytek |
| PE-Dazzle 594 | Isotype | Granzyme B | Granzyme B  Clone: QA16A02 Cat# 372216  Isotype Mouse IgG1  Clone: MOPC-21 Cat# 400176 | BioLegend |
| PE-Fire 640 | CD19 | CD19 | Clone: HIB19 Cat# 302274 | BioLegend |
| PE-Cy5 | – | CD107a | Clone: H4A3 Cat# 555802 | BD Biosciences |
| PE-Fire 700 | CD25 | CD25 | Clone: M-A251 Cat# 356145 | BioLegend |
| PE-Cy7 | – | CD122 | Clone: CF1 Cat# A53365 | Beckman Coulter |
| BUV395 | CD11b | CD11b | Clone: ICRF44 Cat# 563839 | BD Biosciences |
| Live/Dead Blue | Fixable LD Blue | Fixable LD Blue | Viability Cat# L34962 | Thermo Fisher Scientific |
| BUV496 | CD16 | CD16 | Clone: 3G8 Cat# 612945 | BD Biosciences |
| BUV563 | – | CD226 | Clone: DX11 Cat# 748429 | BD Biosciences |
| BUV615 | – | CD159a | Clone: 131411 Cat# 752302 | BD Biosciences |
| BUV661 | CD11c | CD11c | Clone: B-ly6 Cat# 612967 | BD Biosciences |
| BUV737 | CD56 | CD56 | Clone: NCAM16.2  Cat# 612766 | BD Biosciences |
| BUV805 | CD14 | CD14 | Clone: M5E2  Cat# 612902 | BD Biosciences |
| BV421 | CD27 | CD27 | Clone: O323  Cat# 302824 | BioLegend |
| Pacific Blue | – | CD57 | Clone: QA17A04  Cat# 393316 | BioLegend |
| BV480 | Isotype | Ki-67 | Ki-67  Clone B56 Cat# 566109  Isotype Mouse IgG1  Clone: X40 Cat# 1197793 | BD Biosciences |
| BV510 | TCRg/d | TCRg/d | Clone: 11F2 Cat# 745026 | BD Biosciences |
| BV570 | CD8 | CD8 | Clone: RPA-T8 Cat# 301038 | BioLegend |
| BV605 | – | CD336 | Clone: p44-8 Cat# 744301 | BD Biosciences |
| BV650 | – | CD279 | Clone: EH12.1 Cat# 744301 | BD Biosciences |
| BV711 | CD95 | CD95 | Clone: DX2 Cat# 563132 | BD Biosciences |
| BV750 | HLA-DR | HLA-DR | Clone: L243 Cat# 307672 | BioLegend |
| BV786 | CD197 | CD197 | Clone: 2-L1-A Cat# 566758 | BD Biosciences |
| BB515 | CD66b | CD66b | Clone: G10F5 Cat# 564679 | BD Biosciences |
| FITC | CD38 | CD38 | Multiclone Cat# CYT-38F2 | Cytognos |
| Alexa 532 | CD45 | CD45 | Clone: HI30 Cat# 58-0459-42 | Thermo Fisher Scientific |
| BB700 | – | CD314 | Clone: 1D11 Cat# 745863 | BD Biosciences |
| PerCP-eF710 | – | CD69 | Clone: FN50 Cat# 46-0699-42 | Thermo Fisher Scientific |
| APC | – | TIGIT | Clone: 741182 Cat# FAB7898A | R&D Systems |
| NIR Spark 685 | CD45RA | CD45RA | Clone: HI100 Cat# 304168 | BioLegend |
| APC-Alexa 700 | CD127 | CD127 | Clone: R34.34 Cat# A71116 | Beckman Coulter |
| APC-H7 | CD3 | CD3 | Clone: SK7 Cat# 560176 | BD Biosciences |

***Supplemental Table 3. Antibody marker panel used for immunofluorescence***

| **Antibodies used for staining samples:** |  |  |  |
| --- | --- | --- | --- |
| **Stain Buffer:** | **Fluor** | **Clone** | **Catalog number** |
| SignalStain® Antibody Diluent | NA | NA | 8112L |
| Normal Rabbit Serum | NA | NA | 10510 |
| Human TruStain FcX™ | NA | NA | 422302 |
| DAPI | NA | NA | 62248 |
| **Round 1** | **Fluor** | **Clone** | **Catalog number** |
| Calreticulin | AF488 | EPR3924 | ab196158 |
| Gasdermin D | AF555 | E7H9G | Custom Conjugate of 36425 |
| CD138 | AF647 | Mi15 | 356524 |
| CD11b | AF750 | D6X1N | Custom Conjugate of 49420 |
| **Round 2** |  |  |  |
| HMGB1 | AF488 | EPR3507 | ab195010 |
| Ki67 | AF555 | D2H10 | 88338S |
| BCMA | AF647 | E6D7B | 29421S |
| CD163 | AF750 | D6U1J | Custom Conjugate of 93498 |
| **Round 3** |  |  |  |
| HLA-DR | AF488 | E9R2Q | 18915S |
| CD3e | AF555 | D7A6E | 57869S |
| CD4 | AF647 | EPR6855 | ab196147 |
| CD8 | AF750 | D8A8Y | 25648S |
| **Round 4** |  |  |  |
| CD20 | AF488 | L26 | 53-0202-82 |
| Granzyme B | AF555 | D6E9W | 29268S |
| CCR7 | AF647 | EPR23192-57 | ab275165 |
| CD45RA | AF750 | HI100 | CLAF750-65108 |
| **Round 5** |  |  |  |
| Alpha tubulin | AF488 | DM1A | 62204 |
| CD40 (Conjugated to CL555) | CL555 | D8W3N | 40868S |
| CD27 | AF647 | M-T271 | 356434 |
| NaKATPase | AF750 | D4Y7E | 19785S |
| **Round 6** |  |  |  |
| Gasdermin E (Conjugated to CL555) | CL555 | E2X7E | 19453S |
| CD11c | AF647 | D3V1E | 42756S |
| CD45 | AF750 | D9M8I | 16529S |
| **Round 7** |  |  |  |
| CD86 | AF647 | IT2.2 | 305416 |

***Supplemental Table 4. Markers utilized for cell phenotype lineage descriptions in multiplex immuno-fluorescence imaging***

‘-1’ indicates required negative expression of the marker, whereas ‘1’ indicates required positive expression of the marker.

| **Phenotype** | **CD138** | **CD11b** | **BCMA** | **CD163** | **HLA-DR** | **CD3e** | **CD4** | **CD8** | **CD20** | **GzmB** | **CCR7** | **CD45RA** | **CD11c** |
| --- | --- | --- | --- | --- | --- | --- | --- | --- | --- | --- | --- | --- | --- |
| **MM/plasma cells (CD138+)** | 1 |  | -1 |  |  |  |  |  |  |  |  |  |  |
| **MM/plasma cells (CD138+ BCMA+)** | 1 |  | 1 |  |  |  |  |  |  |  |  |  |  |
| **MM/plasma cells (BCMA+)** | -1 |  | 1 |  |  |  |  |  |  |  |  |  |  |
| **B cells** | -1 |  |  |  |  | -1 | -1 | -1 | 1 |  |  |  |  |
| **DCs (CD11b- CD11c+)** |  | -1 |  | -1 | 1 | -1 |  |  | -1 |  |  |  | 1 |
| **Macrophages CD11b-** |  | -1 |  | 1 |  | -1 |  |  | -1 |  |  |  |  |
| **Macrophages CD11b+** |  | 1 |  | 1 |  | -1 |  |  | -1 |  |  |  |  |
| **Monocytes/**  **Granulocytes** |  | 1 |  | -1 |  | -1 |  |  | -1 |  |  |  |  |
| **T cells (DN ie gd T cells)** |  |  |  | -1 |  | 1 | -1 | -1 | -1 |  |  |  |  |
| **T cells (CD4)** |  |  |  | -1 |  | 1 | 1 |  | -1 |  |  |  |  |
| **T cells (CD8)** |  |  |  | -1 |  | 1 |  | 1 | -1 |  |  |  |  |
| **T cells (CD4 central memory)** |  |  |  | -1 |  | 1 | 1 |  | -1 |  | 1 | -1 |  |
| **T cells (CD4 TEMRA)** |  |  |  | -1 |  | 1 | 1 |  | -1 |  | -1 | 1 |  |
| **T cells (CD4 effector memory)** |  |  |  | -1 |  | 1 | 1 |  | -1 |  | -1 | -1 |  |
| **T cells (CD4 naive)** |  |  |  | -1 |  | 1 | 1 |  | -1 |  | 1 | 1 |  |
| **T cells (CD8 central memory)** |  |  |  | -1 |  | 1 |  | 1 | -1 |  | 1 | -1 |  |
| **T cells (CD8 TEMRA)** |  |  |  | -1 |  | 1 |  | 1 | -1 |  | -1 | 1 |  |
| **T cells (CD8 effector memory)** |  |  |  | -1 |  | 1 |  | 1 | -1 |  | -1 | -1 |  |
| **T cells (CD8 naive)** |  |  |  | -1 |  | 1 |  | 1 | -1 |  | 1 | 1 |  |
| **Negative for all makers** | -1 | -1 | -1 | -1 | -1 | -1 | -1 | -1 | -1 |  |  |  | -1 |

***Supplemental Table 5. Tailored mass cytometry panel for immune TME analysis***

| **Target** | **Label** | **Clone** | **Supplier** | **Product ID** |
| --- | --- | --- | --- | --- |
| CD45 | 89Y | HI30 | Standard BioTools | 3089003B |
| CD33 | 111Cd | WM53 | Standard BioTools | 92J042111 |
| CD3 | 141Pr | UCHT1 | Standard BioTools | 3141019B |
| CD19 | 142Nd | HIB19 | Standard BioTools | 3142001B |
| CD5 | 143Nd | UCHT2 | Standard BioTools | 3143007B |
| CD1c | 144Nd | L161 | BioLegend | 331502 |
| CD226 | 145Nd | 11A8 | BioLegend | 338302 |
| CD8a | 146Nd | RPA-T8 | Standard BioTools | 3146001B |
| CD11c | 147Sm | Bu15 | Standard BioTools | 3147008B |
| CD16 | 148Nd | 3G8 | Standard BioTools | 3148004B |
| CD127 | 149Sm | A019D5 | Standard BioTools | 3149011B |
| CD138 | 150Nd | DL101 | Standard BioTools | 3150012B |
| CD123 | 151Eu | 6H6 | Standard BioTools | 3151001B |
| NKG2A | 152Sm | REA110 | Miltenyi | 130-122-329 |
| TIGIT | 153Eu | MBSA43 | Standard BioTools | 3153019B |
| TIM-3 | 154Sm | F38-2E2 | Standard BioTools | 3154010B |
| CD45RA | 155Gd | HI100 | Standard BioTools | 3155011B |
| PD-L1 | 156Gd | 29E.2A3 | Standard BioTools | 3156026B |
| CD27 | 158Gd | L128 | Standard BioTools | 3158010B |
| CCR7 | 159Tb | G043H7 | Standard BioTools | 3159003A |
| CD28 | 160Gd | CD28.2 | Standard BioTools | 3160003B |
| CD66B | 162Dy | 80H3 | Standard BioTools | 3162023B |
| CXCR3 | 163Dy | G025H7 | Standard BioTools | 3163004B |
| KLRG1 | 164Dy | 2388L | R&D | MAB70293 |
| CD43 | 165Ho | 290111 | R&D | MAB2038 |
| NKG2D | 166Er | ON72 | Standard BioTools | 3166016B |
| CD38 | 167Er | HIT2 | Standard BioTools | 3167001B |
| ICOS | 168Er | C398.4A | Standard BioTools | 3168024B |
| CD25 | 169Tm | 2A3 | Standard BioTools | 3169003B |
| HLA-DR | 170Er | K243 | Standard BioTools | 3170013B |
| CD4 | 171Yb | RPA-T4 | BioLegend | 300502 |
| CD57 | 172Yb | HCD57 | Standard BioTools | 3172009B |
| PD-1 | 174Yb | EH12.ZH7 | Standard BioTools | 3174020B |
| CD14 | 175Lu | M5E2 | Standard BioTools | 3175015B |
| CD56 | 176Yb | NCAM16.2 | Standard BioTools | 3176008B |
| CD11b | 209Bi | ICRF44 | Standard BioTools | 3209003B |
| T-bet | 113In | 4B10 | BioLegend | 644802 |
| Ki67 | 161Dy | B56 | Standard BioTools | 3161007B |
| Granzyme B | 173Yb | GB11 | Standard BioTools | 3173006B |

***Supplemental Table 6. Response categories for DREAMM-14 serum analysis***

| Response category | Number of patients in response category | Best confirmed response | Best confirmed response N |
| --- | --- | --- | --- |
| Deep responder | 24 | Stringent complete response | 7 |
| Deep responder | 24 | Complete response | 5 |
| Deep responder | 24 | Very good partial response | 12 |
| Modest responder | 11 | Partial response | 4 |
| Modest responder | 11 | Minimal response | 3 |
| Modest responder | 11 | Stable disease | 4 |
| Non responder | 7 | Progressive disease | 7 |

***Supplemental File 1. Custom marker panel for spatial transcriptomics***

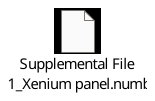

***Supplemental File 2. Olink proteomics data***

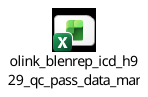
